## Supplementary material for "Real-time Forecasting of Data Revisions in Epidemic Surveillance Streams": S1 Appendix

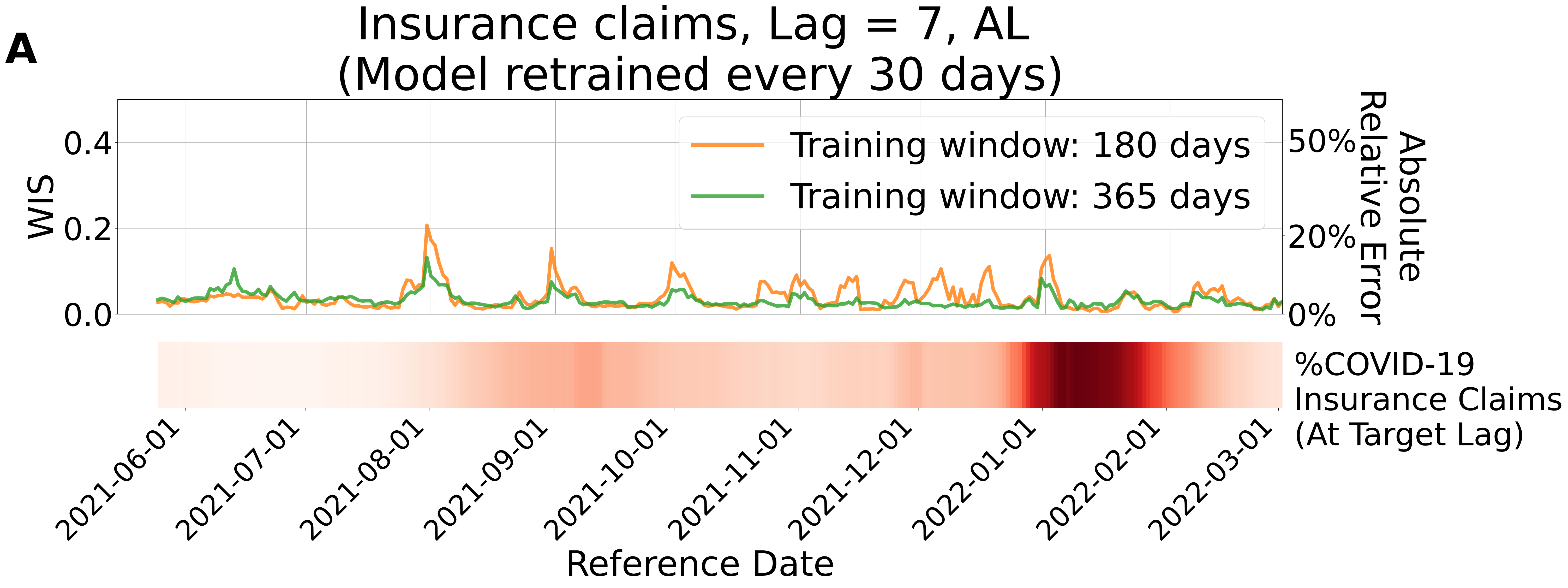
A
Insurance claims, Lag = 7, AL
(Model retrained every 30 days)
WIS
0.0
0.2
0.4
Absolute Relative Error
0%
20%
50%
Training window: 180 days
Training window: 365 days
%COVID-19 Insurance Claims (At Target Lag)
2021-06-01
2021-07-01
2021-08-01
2021-09-01
2021-10-01
2021-11-01
2021-12-01
2022-01-01
2022-02-01
2022-03-01
Reference Date

B

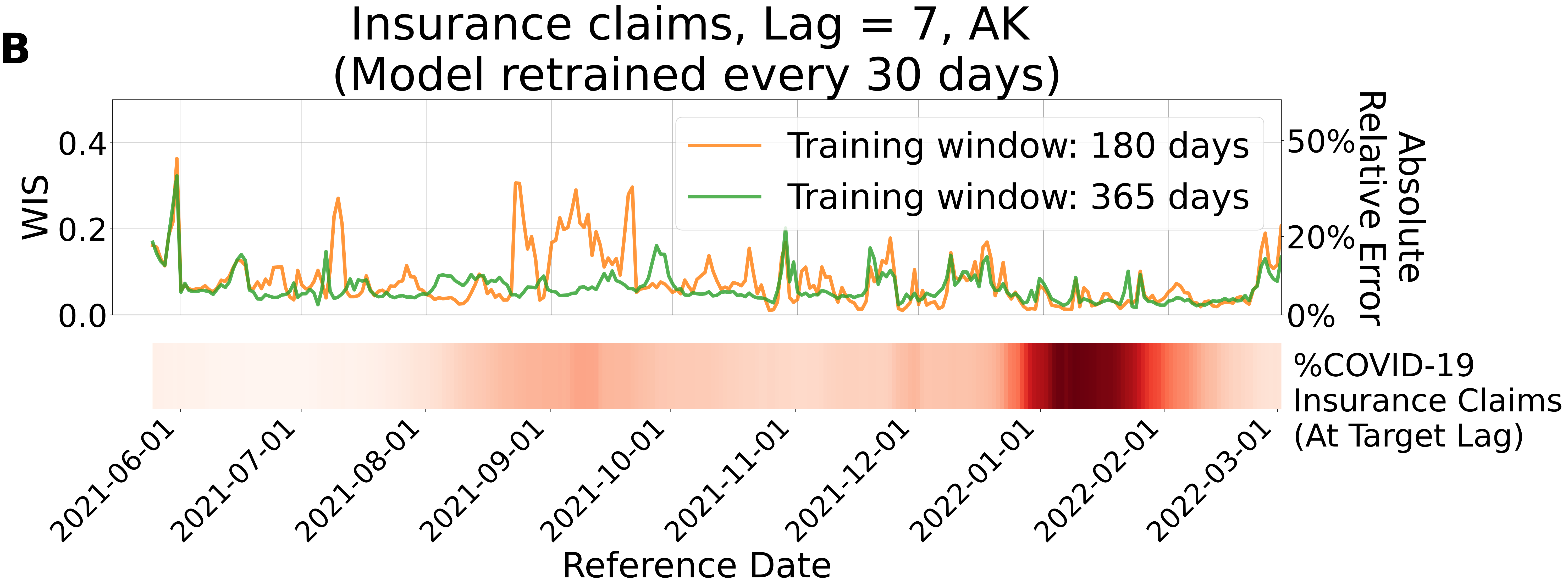

Insurance claims, Lag = 7, AK
(Model retrained every 30 days)
WIS
0.4
0.2
0.0
Absolute Relative Error
50%
20%
0%
Training window: 180 days
Training window: 365 days
%COVID-19
Insurance Claims
(At Target Lag)
2021-06-01
2021-07-01
2021-08-01
2021-09-01
2021-10-01
2021-11-01
2021-12-01
2022-01-01
2022-02-01
2022-03-01
Reference Date

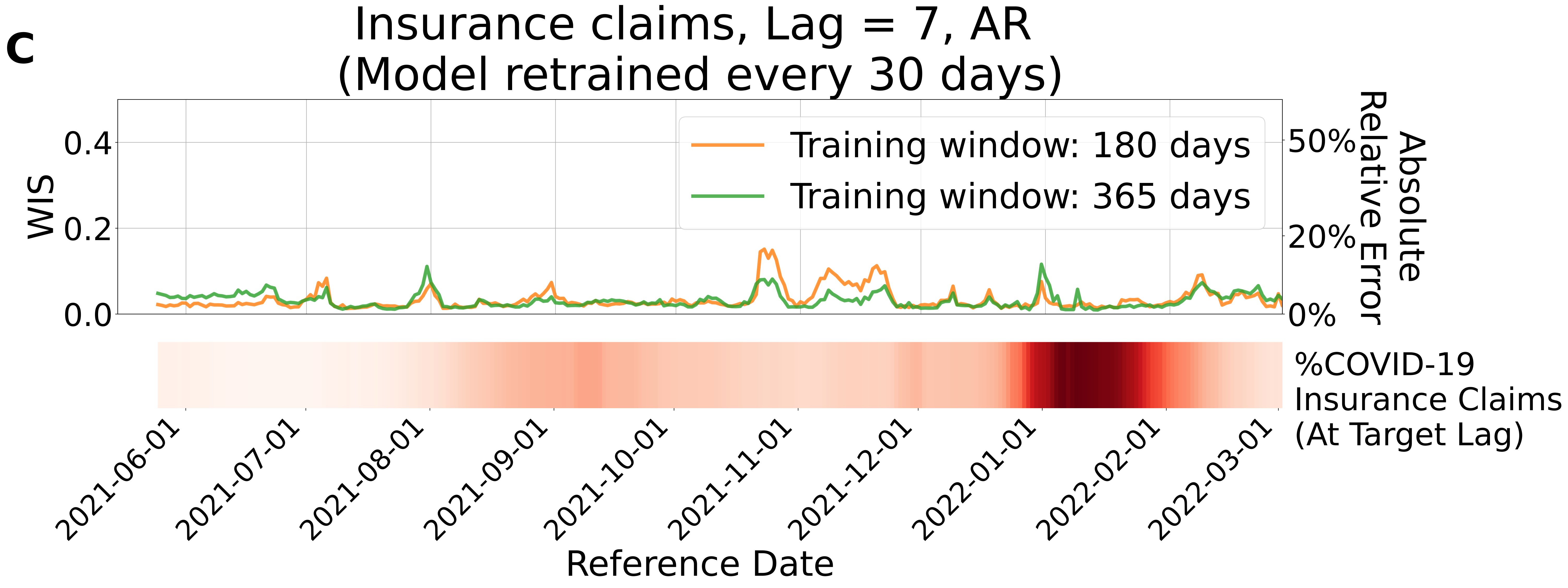
c
Insurance claims, Lag = 7, AR
(Model retrained every 30 days)
Training window: 180 days
Training window: 365 days
WIS
0.0
0.2
0.4
Absolute Relative Error
0%
20%
50%
%COVID-19 Insurance Claims (At Target Lag)
2021-06-01
2021-07-01
2021-08-01
2021-09-01
2021-10-01
2021-11-01
2021-12-01
2022-01-01
2022-02-01
2022-03-01
Reference Date

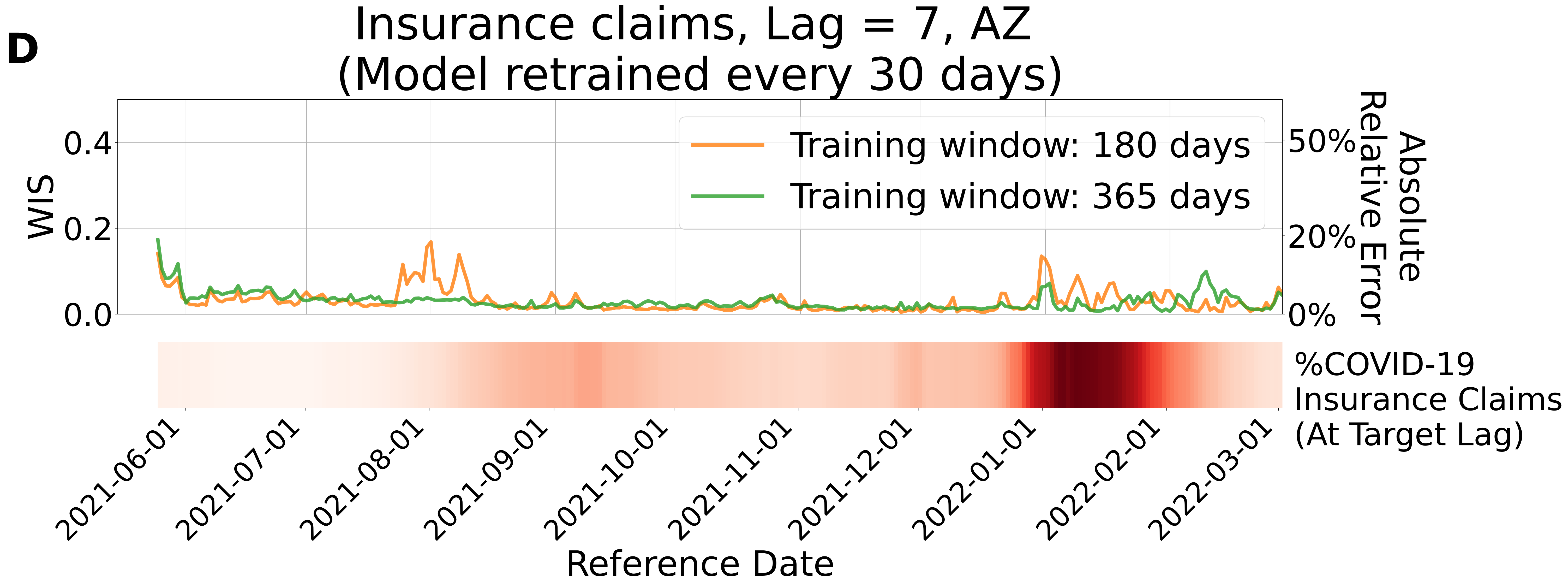

D
Insurance claims, Lag = 7, AZ
(Model retrained every 30 days)
WIS
0.0
0.2
0.4
Absolute Relative Error
0%
20%
50%
Training window: 180 days
Training window: 365 days
%COVID-19
Insurance Claims
(At Target Lag)
2021-06-01
2021-07-01
2021-08-01
2021-09-01
2021-10-01
2021-11-01
2021-12-01
2022-01-01
2022-02-01
2022-03-01
Reference Date

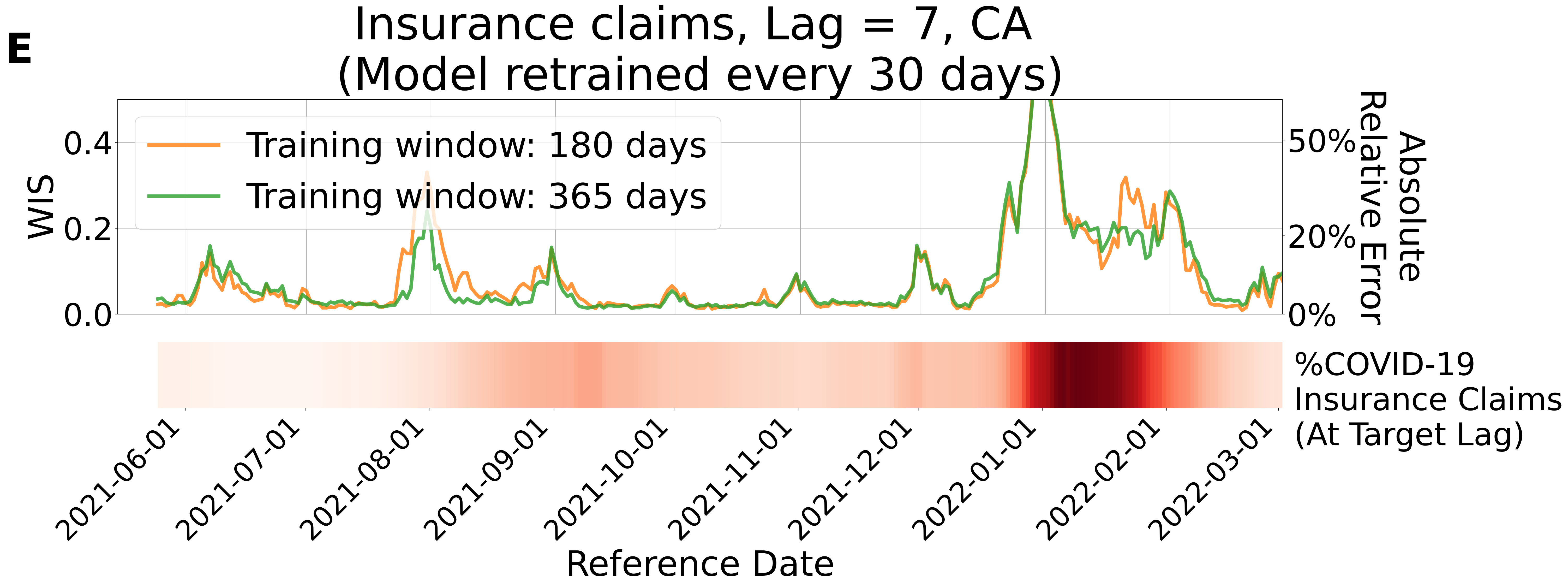
E
Insurance claims, Lag = 7, CA
(Model retrained every 30 days)
Training window: 180 days
Training window: 365 days
WIS
0.4
0.2
0.0
Absolute Relative Error
50%
20%
0%
%COVID-19
Insurance Claims
(At Target Lag)
2021-06-01
2021-07-01
2021-08-01
2021-09-01
2021-10-01
2021-11-01
2021-12-01
2022-01-01
2022-02-01
2022-03-01
Reference Date

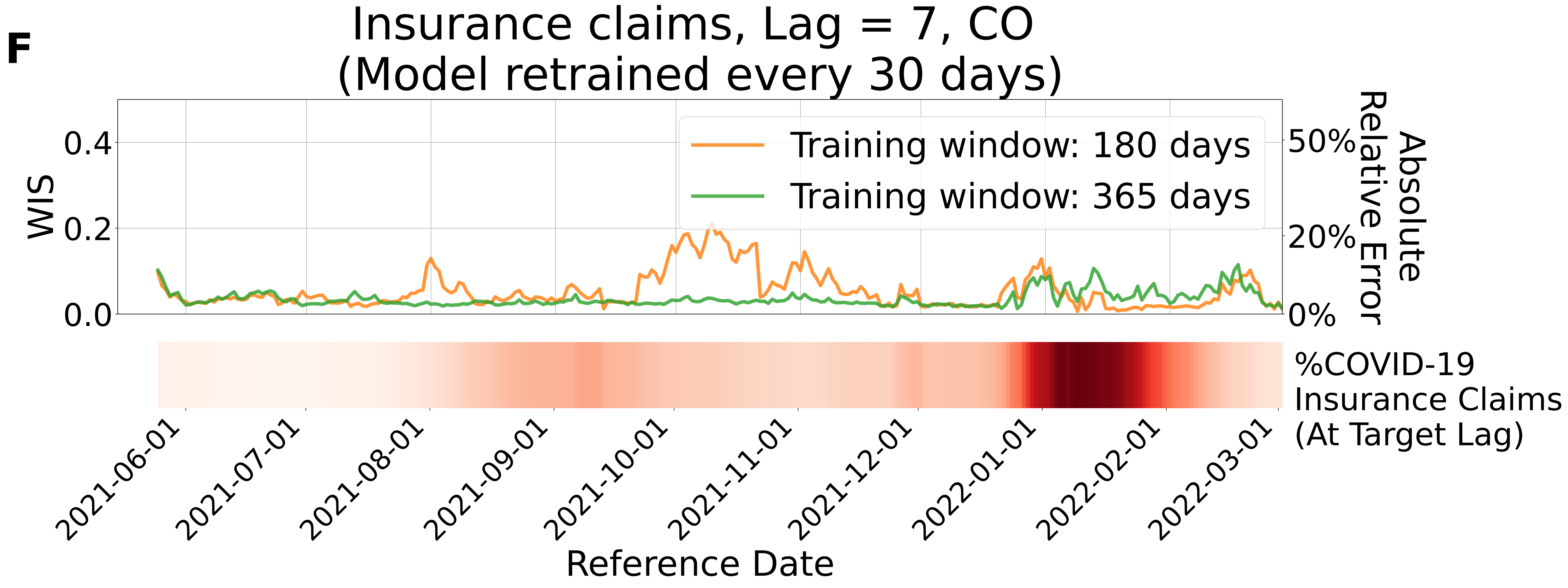
F
Insurance claims, Lag = 7, CO
(Model retrained every 30 days)
WIS
0.0
0.2
0.4
Absolute Relative Error
0%
20%
50%
Training window: 180 days
Training window: 365 days
%COVID-19 Insurance Claims (At Target Lag)
2021-06-01
2021-07-01
2021-08-01
2021-09-01
2021-10-01
2021-11-01
2021-12-01
2022-01-01
2022-02-01
2022-03-01
Reference Date

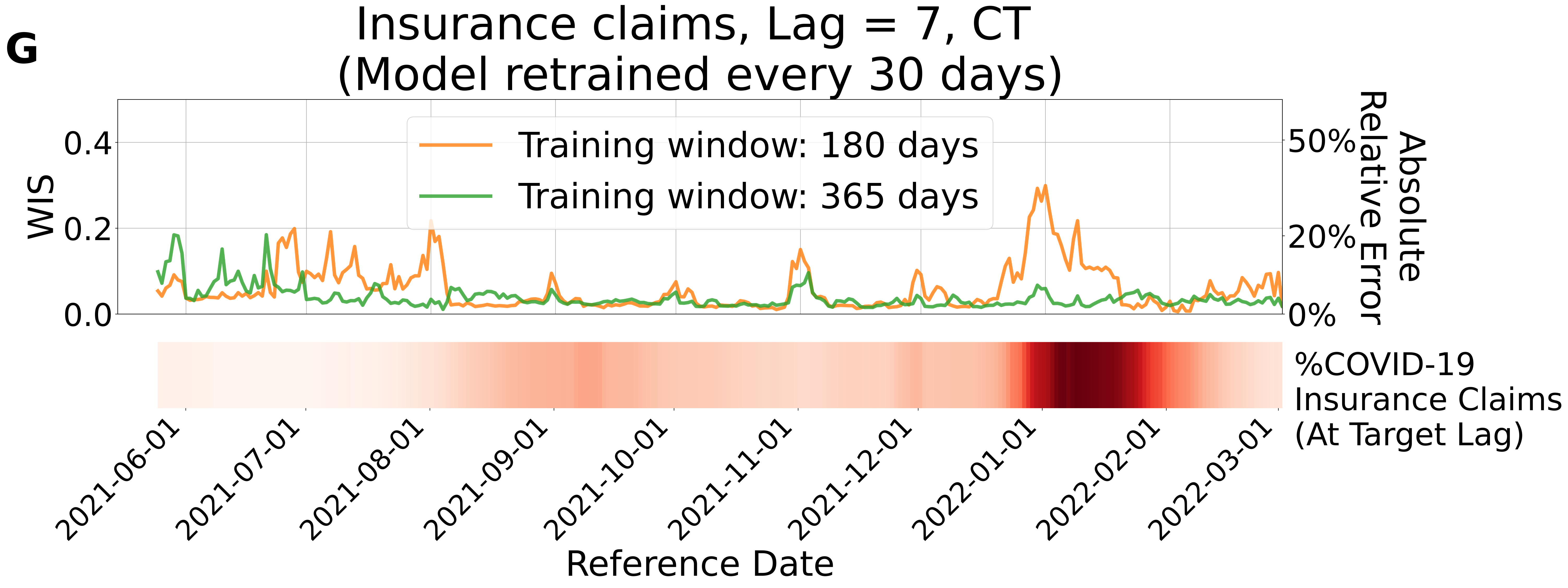
G
Insurance claims, Lag = 7, CT
(Model retrained every 30 days)
Training window: 180 days
Training window: 365 days
WIS
0.0
0.2
0.4
Absolute Relative Error
0%
20%
50%
%COVID-19 Insurance Claims (At Target Lag)
2021-06-01
2021-07-01
2021-08-01
2021-09-01
2021-10-01
2021-11-01
2021-12-01
2022-01-01
2022-02-01
2022-03-01
Reference Date

**H**

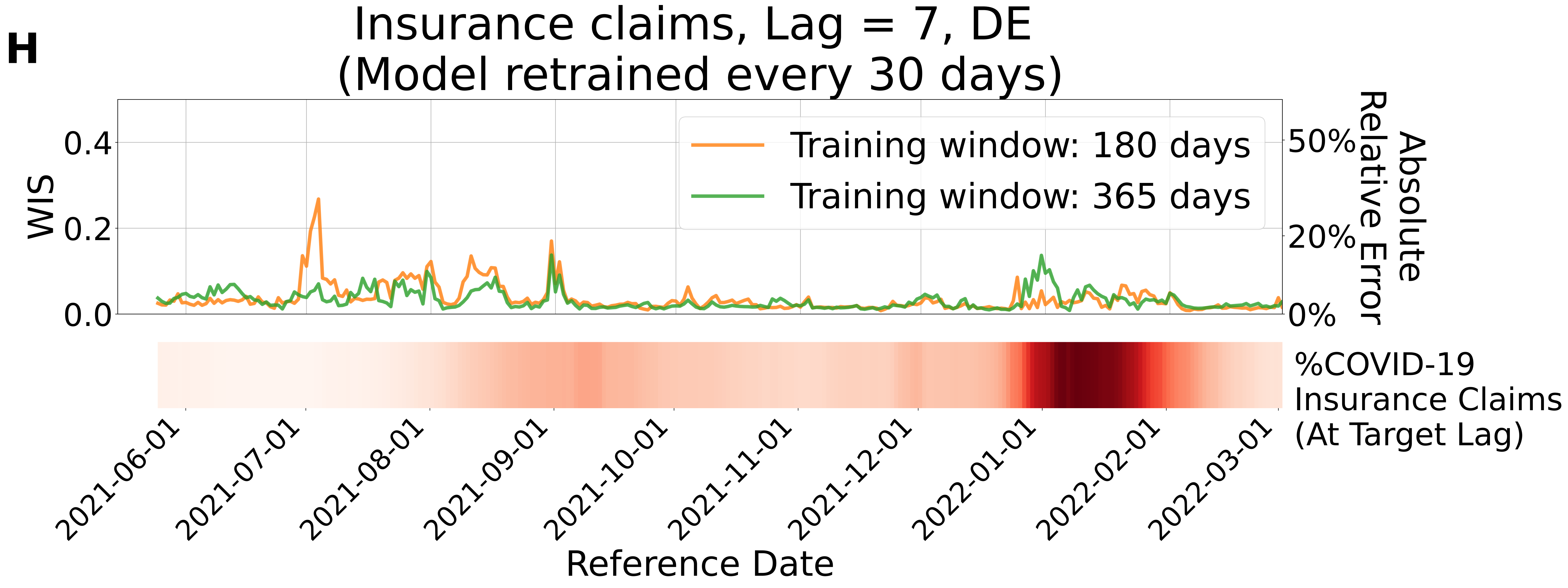

Insurance claims, Lag = 7, DE
(Model retrained every 30 days)
WIS
0.0
0.2
0.4
Absolute Relative Error
0%
20%
50%
Training window: 180 days
Training window: 365 days
%COVID-19 Insurance Claims (At Target Lag)
2021-06-01
2021-07-01
2021-08-01
2021-09-01
2021-10-01
2021-11-01
2021-12-01
2022-01-01
2022-02-01
2022-03-01
Reference Date

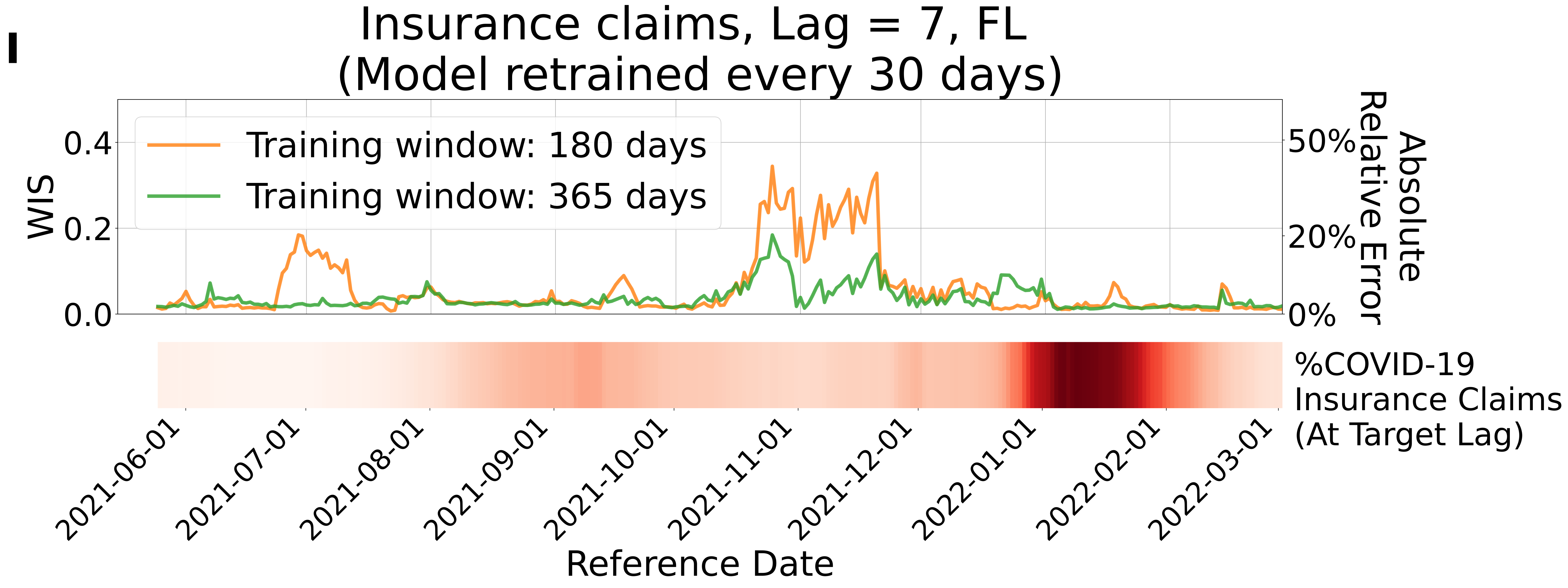

I
Insurance claims, Lag = 7, FL
(Model retrained every 30 days)
Training window: 180 days
Training window: 365 days
WIS
0.0
0.2
0.4
Absolute Relative Error
0%
20%
50%
%COVID-19 Insurance Claims (At Target Lag)
2021-06-01
2021-07-01
2021-08-01
2021-09-01
2021-10-01
2021-11-01
2021-12-01
2022-01-01
2022-02-01
2022-03-01
Reference Date

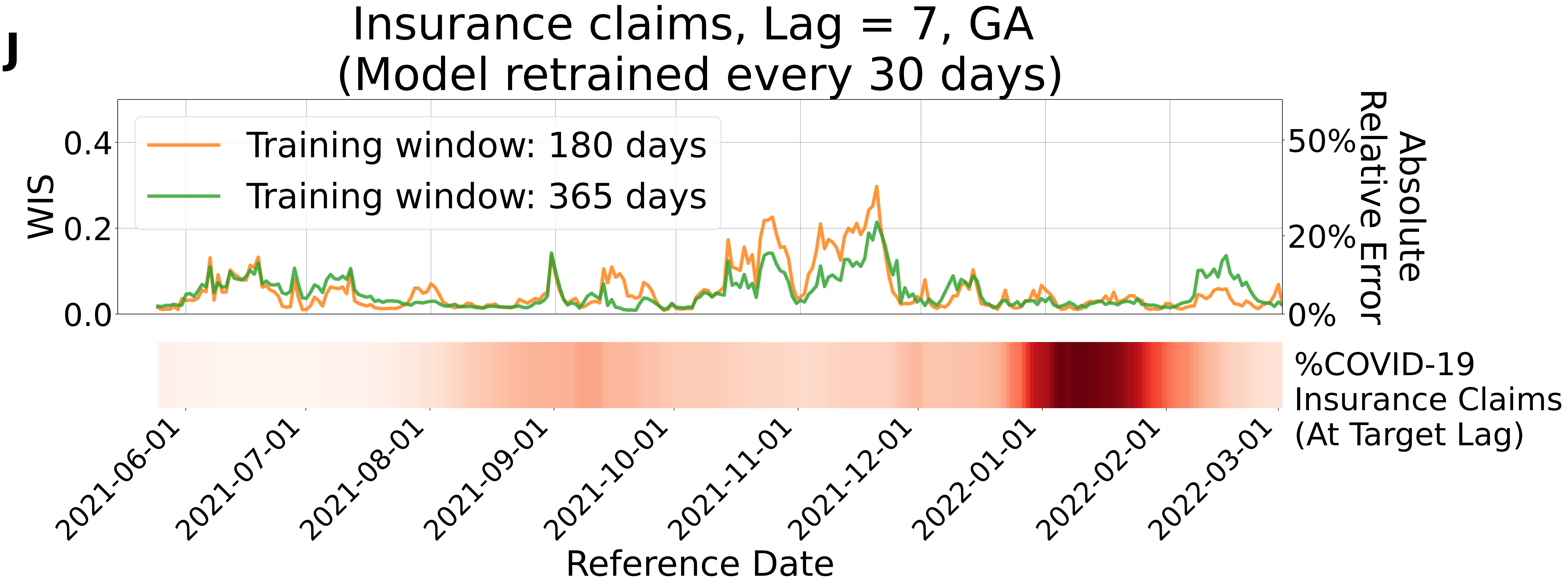
J
Insurance claims, Lag = 7, GA
(Model retrained every 30 days)
Training window: 180 days
Training window: 365 days
WIS
0.0
0.2
0.4
Absolute Relative Error
0%
20%
50%
%COVID-19 Insurance Claims (At Target Lag)
2021-06-01
2021-07-01
2021-08-01
2021-09-01
2021-10-01
2021-11-01
2021-12-01
2022-01-01
2022-02-01
2022-03-01
Reference Date

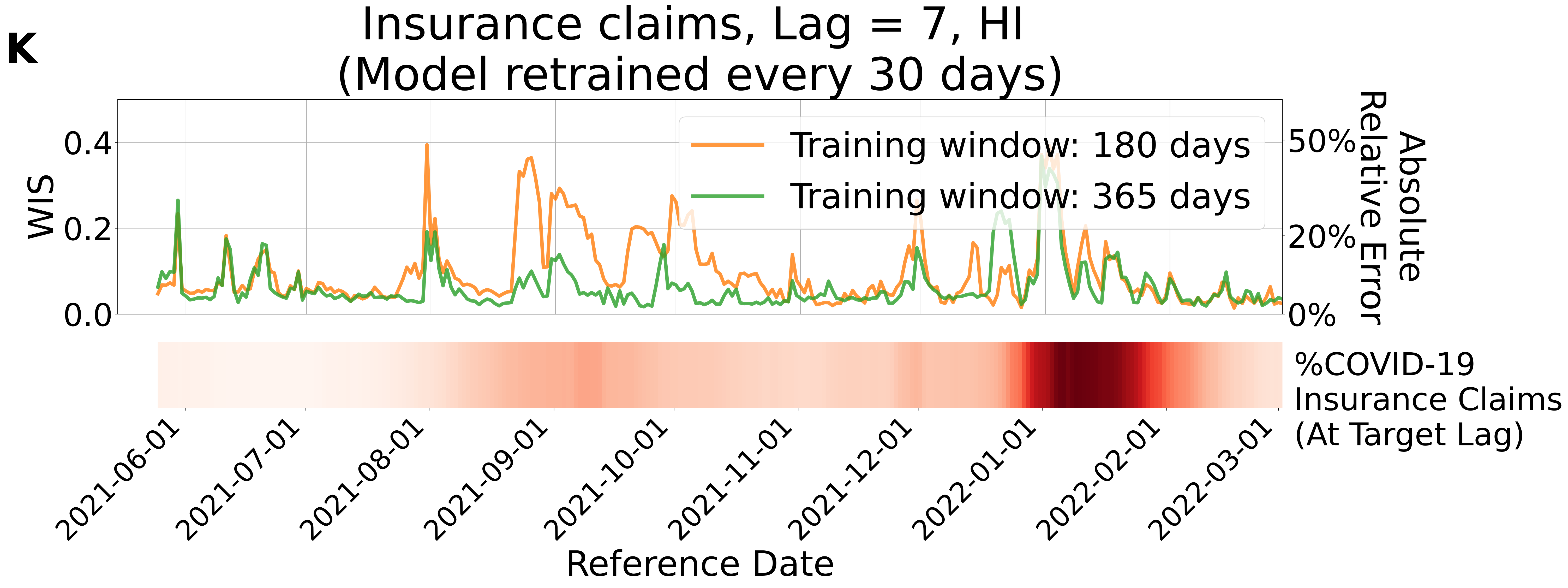
K
Insurance claims, Lag = 7, HI
(Model retrained every 30 days)
WIS
0.0
0.2
0.4
Absolute Relative Error
0%
20%
50%
Training window: 180 days
Training window: 365 days
%COVID-19
Insurance Claims
(At Target Lag)
2021-06-01
2021-07-01
2021-08-01
2021-09-01
2021-10-01
2021-11-01
2021-12-01
2022-01-01
2022-02-01
2022-03-01
Reference Date

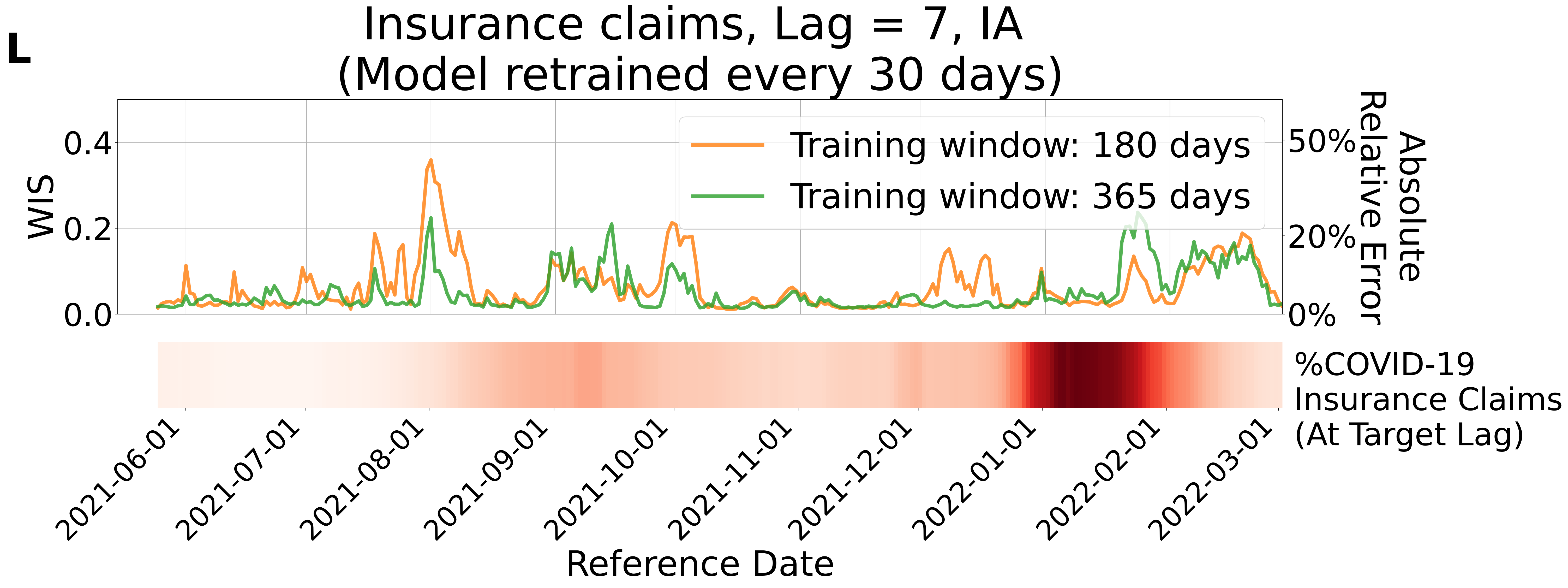

L
Insurance claims, Lag = 7, IA
(Model retrained every 30 days)
WIS
0.0
0.2
0.4
Absolute Relative Error
0%
20%
50%
Training window: 180 days
Training window: 365 days
%COVID-19 Insurance Claims (At Target Lag)
2021-06-01
2021-07-01
2021-08-01
2021-09-01
2021-10-01
2021-11-01
2021-12-01
2022-01-01
2022-02-01
2022-03-01
Reference Date

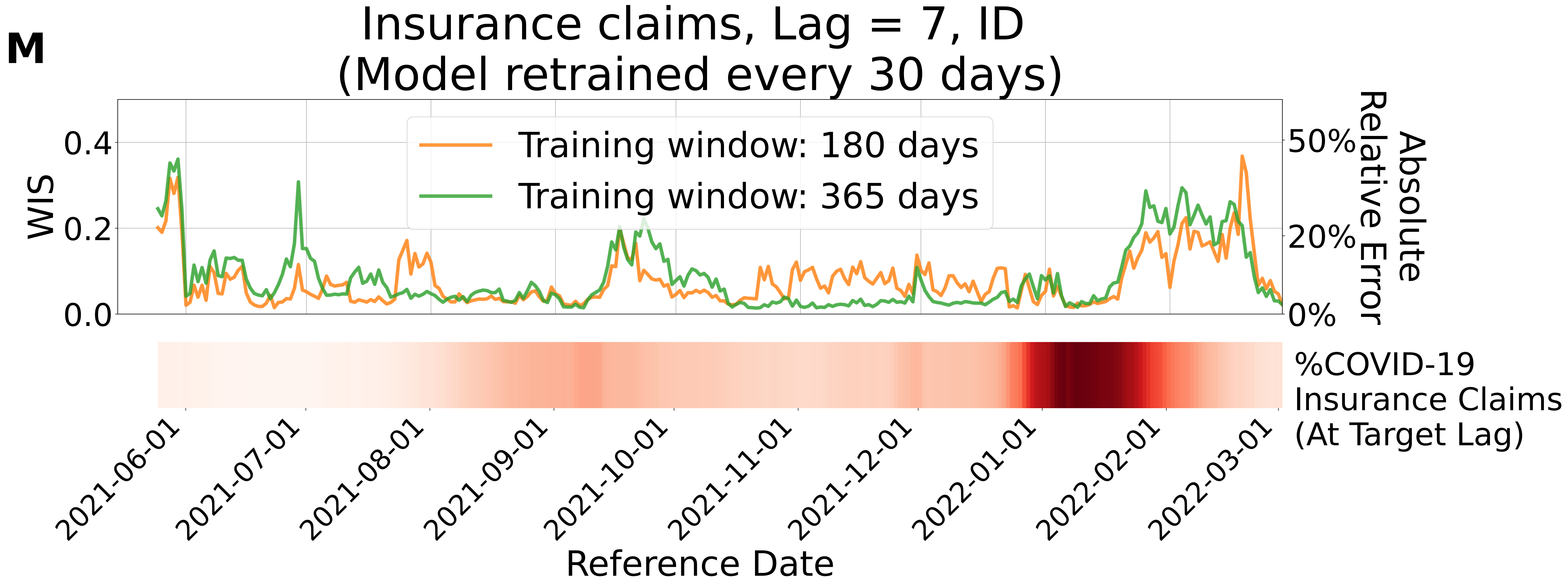

M
Insurance claims, Lag = 7, ID
(Model retrained every 30 days)
Training window: 180 days
Training window: 365 days
WIS
0.0
0.2
0.4
Absolute Relative Error
0%
20%
50%
%COVID-19 Insurance Claims (At Target Lag)
2021-06-01
2021-07-01
2021-08-01
2021-09-01
2021-10-01
2021-11-01
2021-12-01
2022-01-01
2022-02-01
2022-03-01
Reference Date

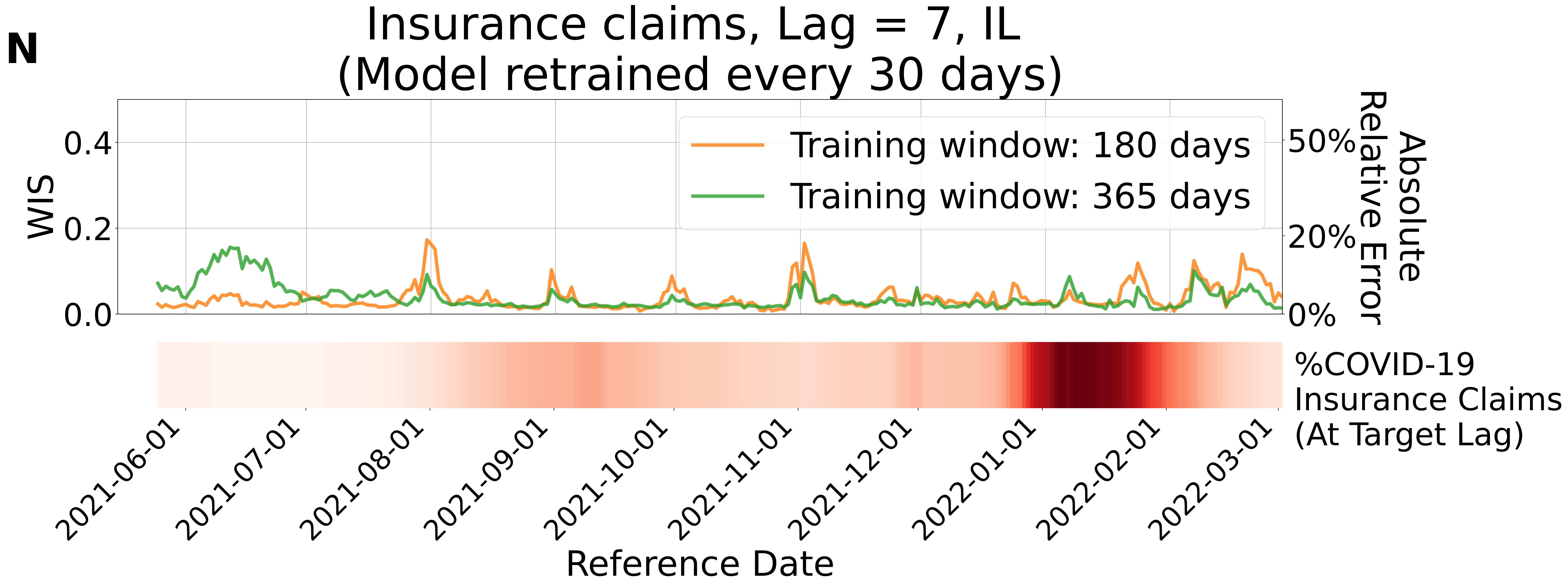
N
Insurance claims, Lag = 7, IL
(Model retrained every 30 days)
WIS
0.0
0.2
0.4
Absolute Relative Error
0%
20%
50%
Training window: 180 days
Training window: 365 days
%COVID-19 Insurance Claims (At Target Lag)
2021-06-01
2021-07-01
2021-08-01
2021-09-01
2021-10-01
2021-11-01
2021-12-01
2022-01-01
2022-02-01
2022-03-01
Reference Date

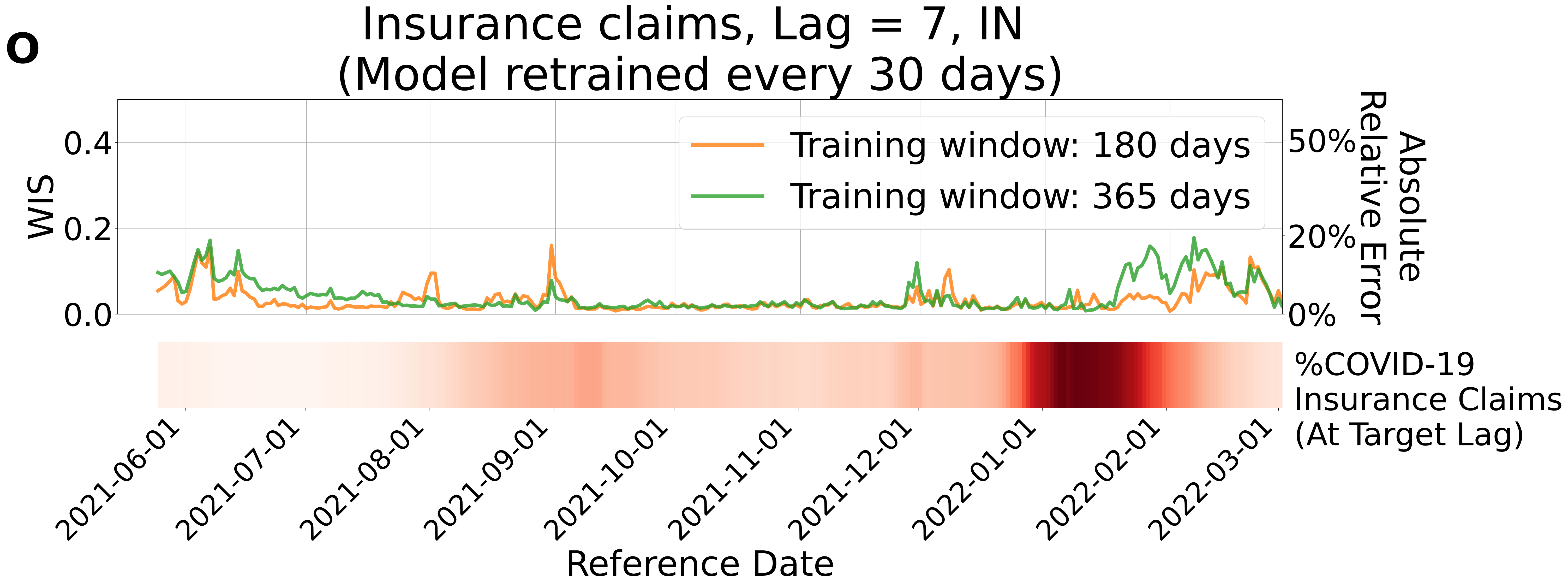
o
Insurance claims, Lag = 7, IN
(Model retrained every 30 days)
Training window: 180 days
Training window: 365 days
WIS
0.4
0.2
0.0
Absolute Relative Error
50%
20%
0%
%COVID-19 Insurance Claims (At Target Lag)
2021-06-01
2021-07-01
2021-08-01
2021-09-01
2021-10-01
2021-11-01
2021-12-01
2022-01-01
2022-02-01
2022-03-01
Reference Date

**P**

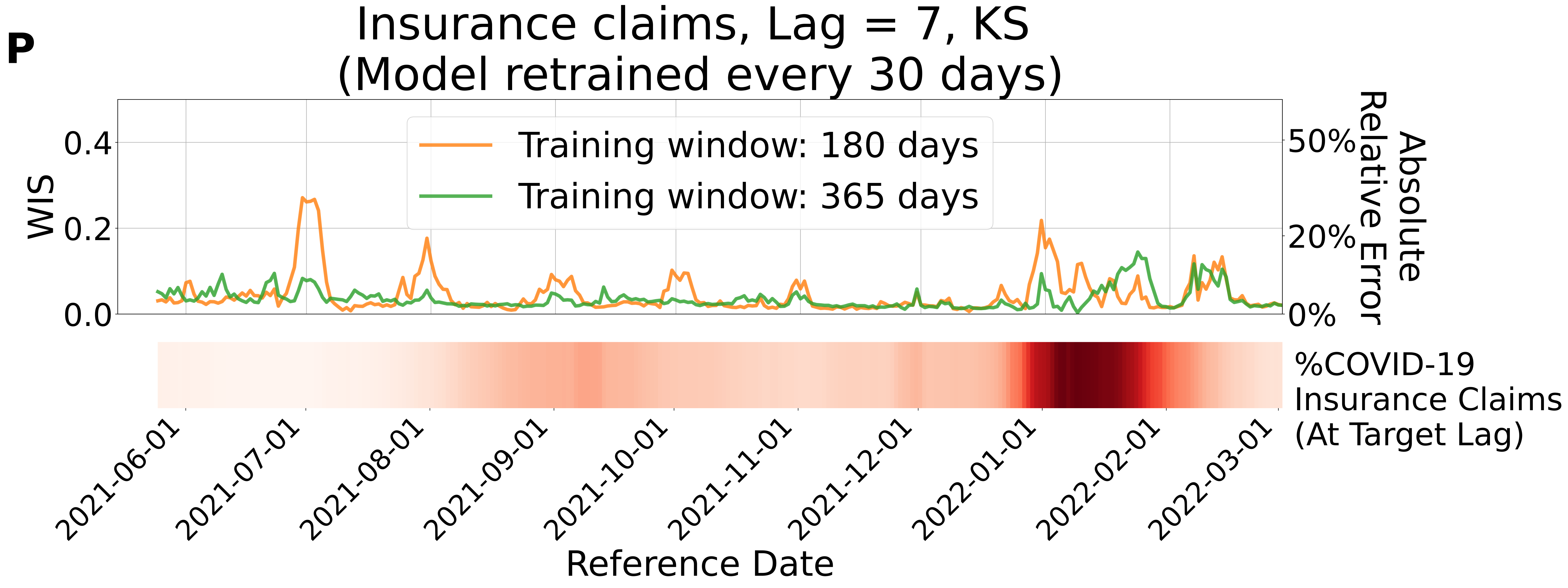

Insurance claims, Lag = 7, KS
(Model retrained every 30 days)
Training window: 180 days
Training window: 365 days
WIS
0.0
0.2
0.4
Absolute Relative Error
0%
20%
50%
%COVID-19 Insurance Claims (At Target Lag)
2021-06-01
2021-07-01
2021-08-01
2021-09-01
2021-10-01
2021-11-01
2021-12-01
2022-01-01
2022-02-01
2022-03-01
Reference Date

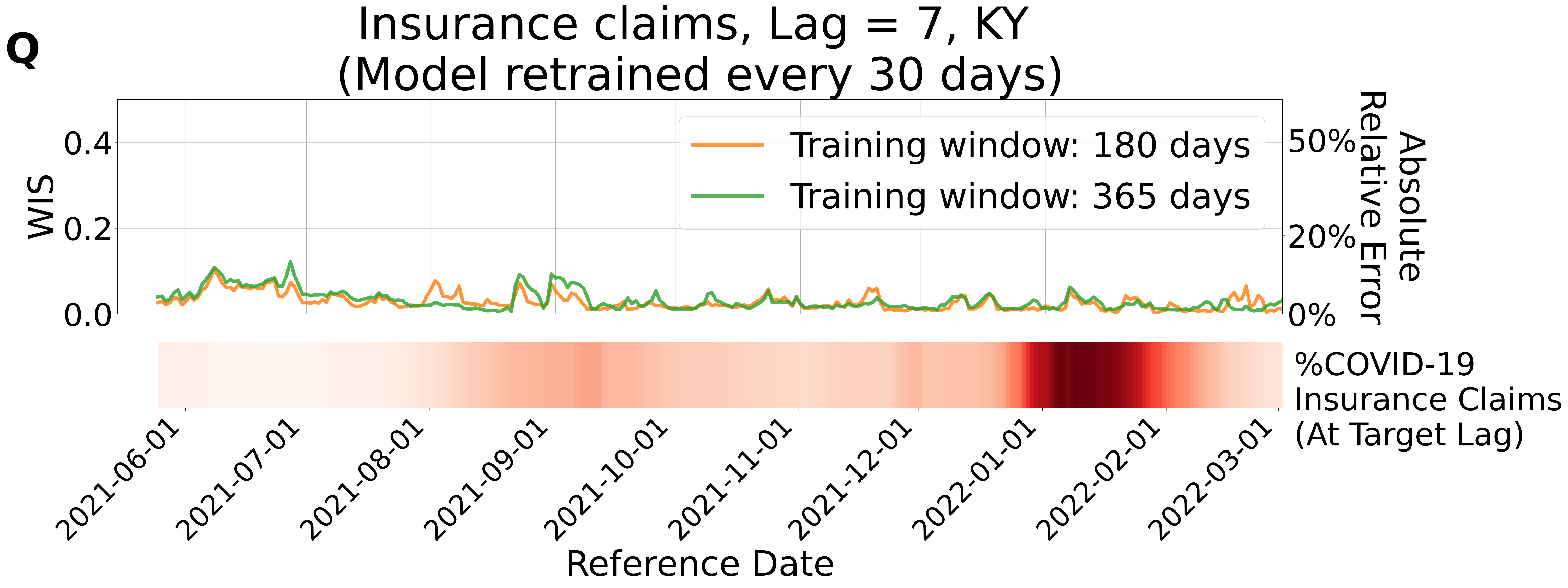
Q
Insurance claims, Lag = 7, KY
(Model retrained every 30 days)
WIS
0.4
0.2
0.0
Training window: 180 days
Training window: 365 days
50%
20%
0%
Absolute
Relative Error
%COVID-19
Insurance Claims
(At Target Lag)
2021-06-01
2021-07-01
2021-08-01
2021-09-01
2021-10-01
2021-11-01
2021-12-01
2022-01-01
2022-02-01
2022-03-01
Reference Date

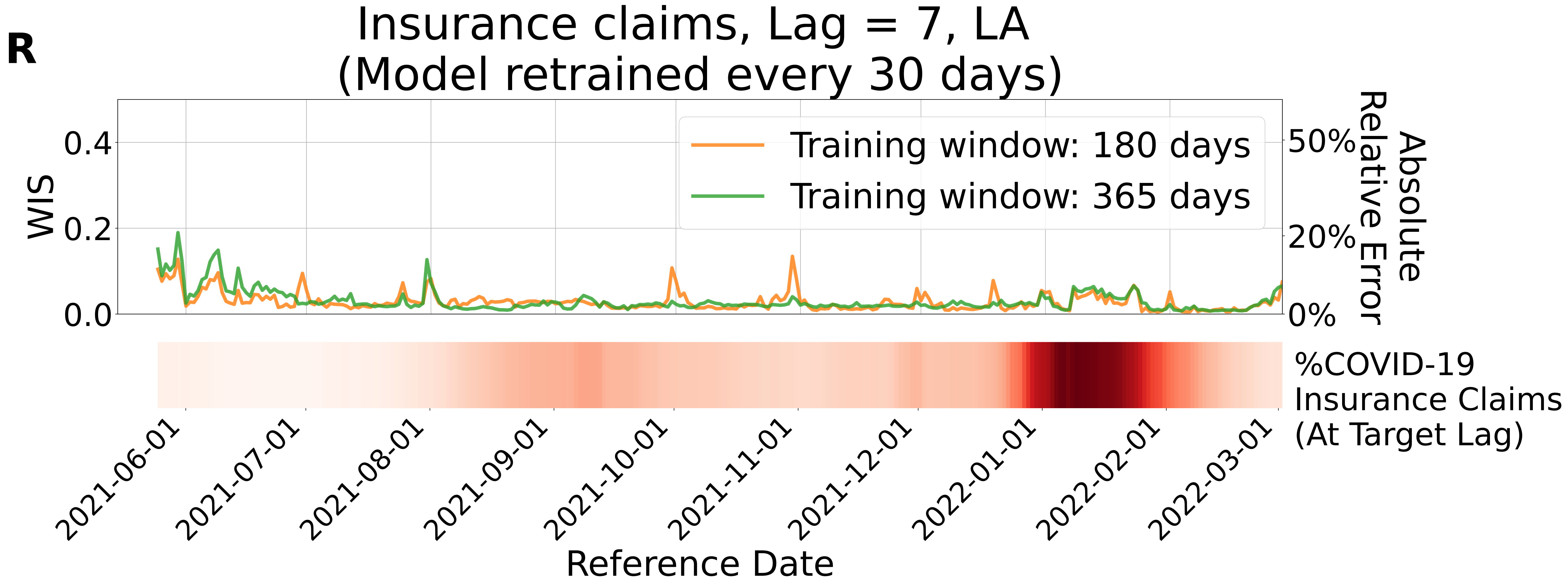
R
Insurance claims, Lag = 7, LA
(Model retrained every 30 days)
WIS
0.0
0.2
0.4
Absolute Relative Error
0%
20%
50%
Training window: 180 days
Training window: 365 days
%COVID-19 Insurance Claims (At Target Lag)
2021-06-01
2021-07-01
2021-08-01
2021-09-01
2021-10-01
2021-11-01
2021-12-01
2022-01-01
2022-02-01
2022-03-01
Reference Date

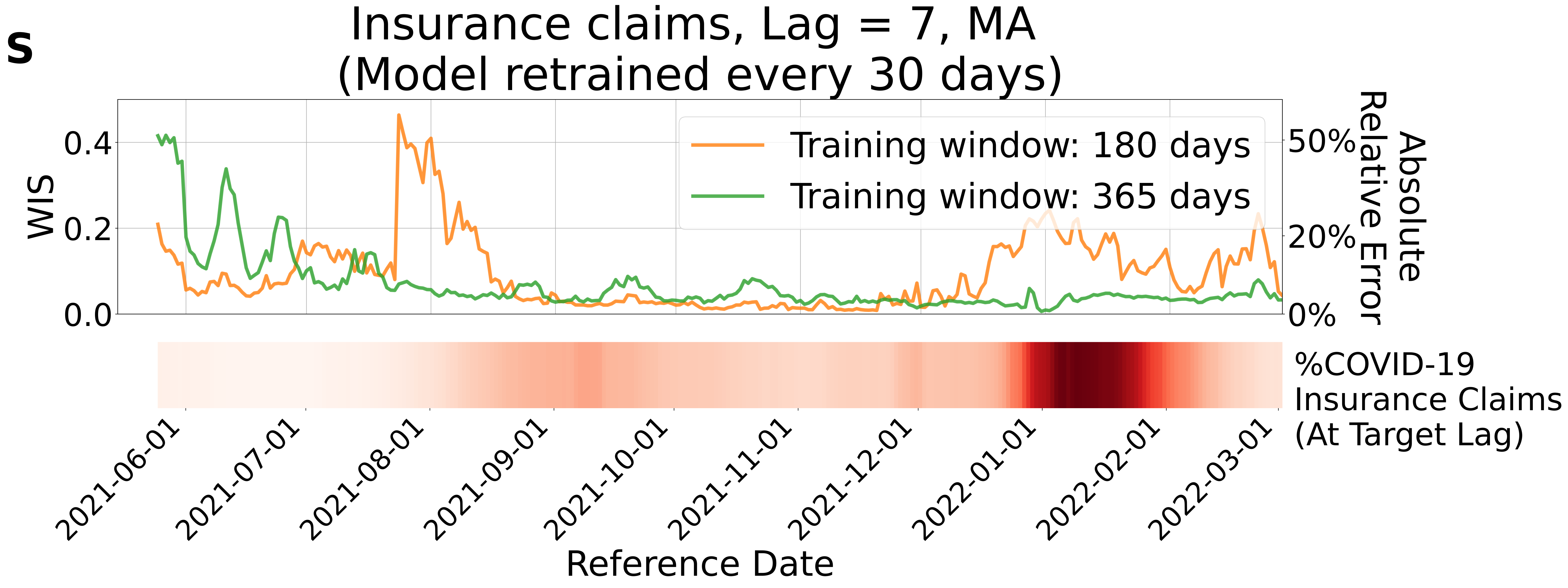
s
Insurance claims, Lag = 7, MA
(Model retrained every 30 days)
WIS
0.0
0.2
0.4
Absolute Relative Error
0%
20%
50%
Training window: 180 days
Training window: 365 days
%COVID-19 Insurance Claims (At Target Lag)
2021-06-01
2021-07-01
2021-08-01
2021-09-01
2021-10-01
2021-11-01
2021-12-01
2022-01-01
2022-02-01
2022-03-01
Reference Date

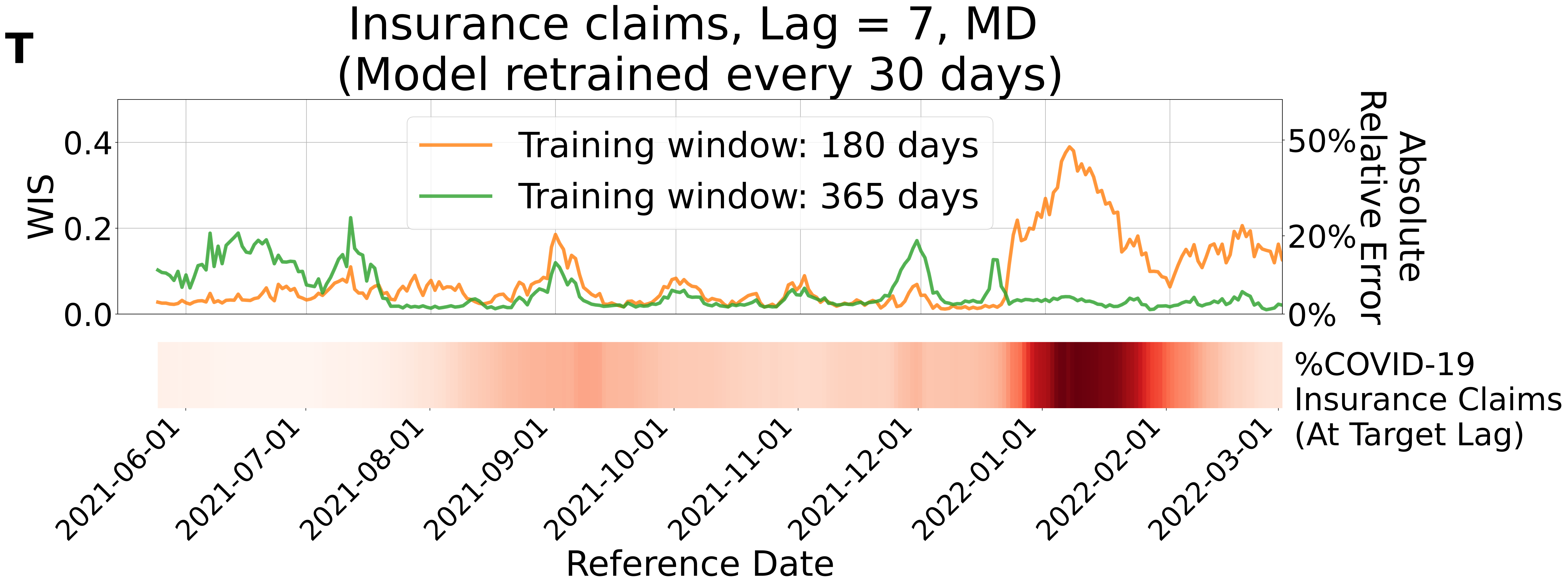

T
Insurance claims, Lag = 7, MD
(Model retrained every 30 days)
Training window: 180 days
Training window: 365 days
WIS
0.0
0.2
0.4
Absolute Relative Error
0%
20%
50%
%COVID-19 Insurance Claims (At Target Lag)
2021-06-01
2021-07-01
2021-08-01
2021-09-01
2021-10-01
2021-11-01
2021-12-01
2022-01-01
2022-02-01
2022-03-01
Reference Date

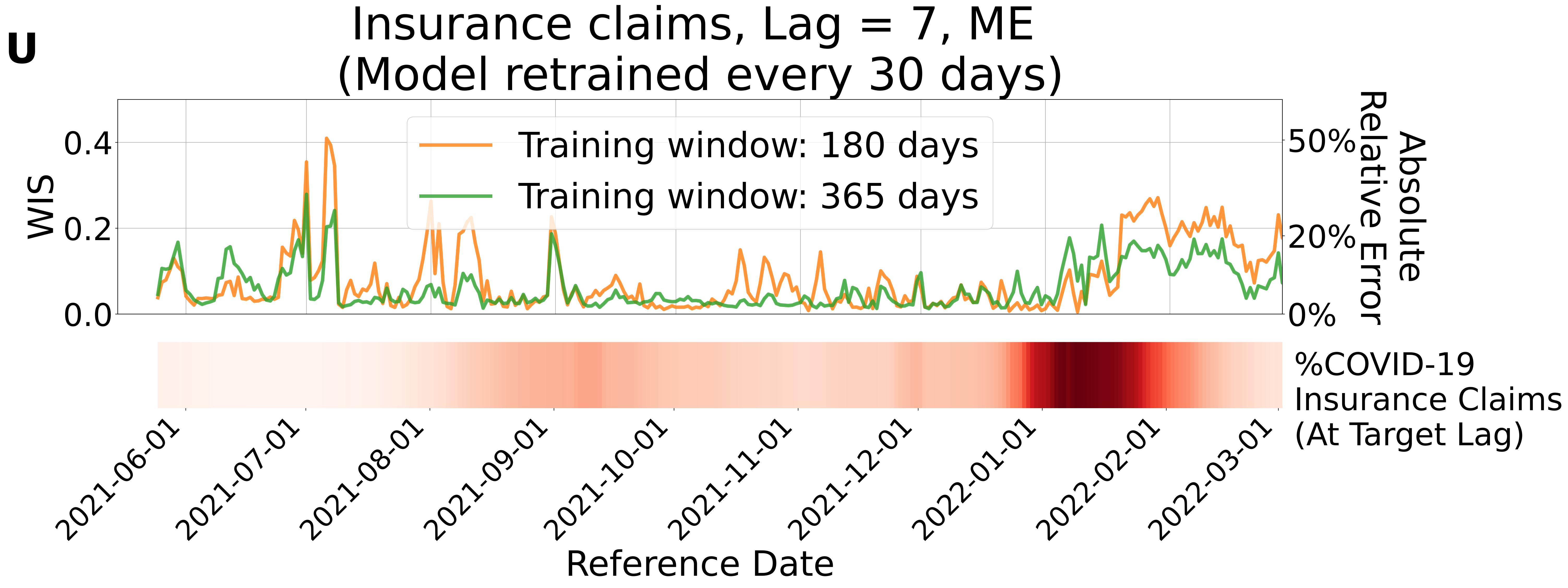
U
Insurance claims, Lag = 7, ME
(Model retrained every 30 days)
Training window: 180 days
Training window: 365 days
WIS
0.0
0.2
0.4
Absolute Relative Error
0%
20%
50%
%COVID-19 Insurance Claims (At Target Lag)
2021-06-01
2021-07-01
2021-08-01
2021-09-01
2021-10-01
2021-11-01
2021-12-01
2022-01-01
2022-02-01
2022-03-01
Reference Date

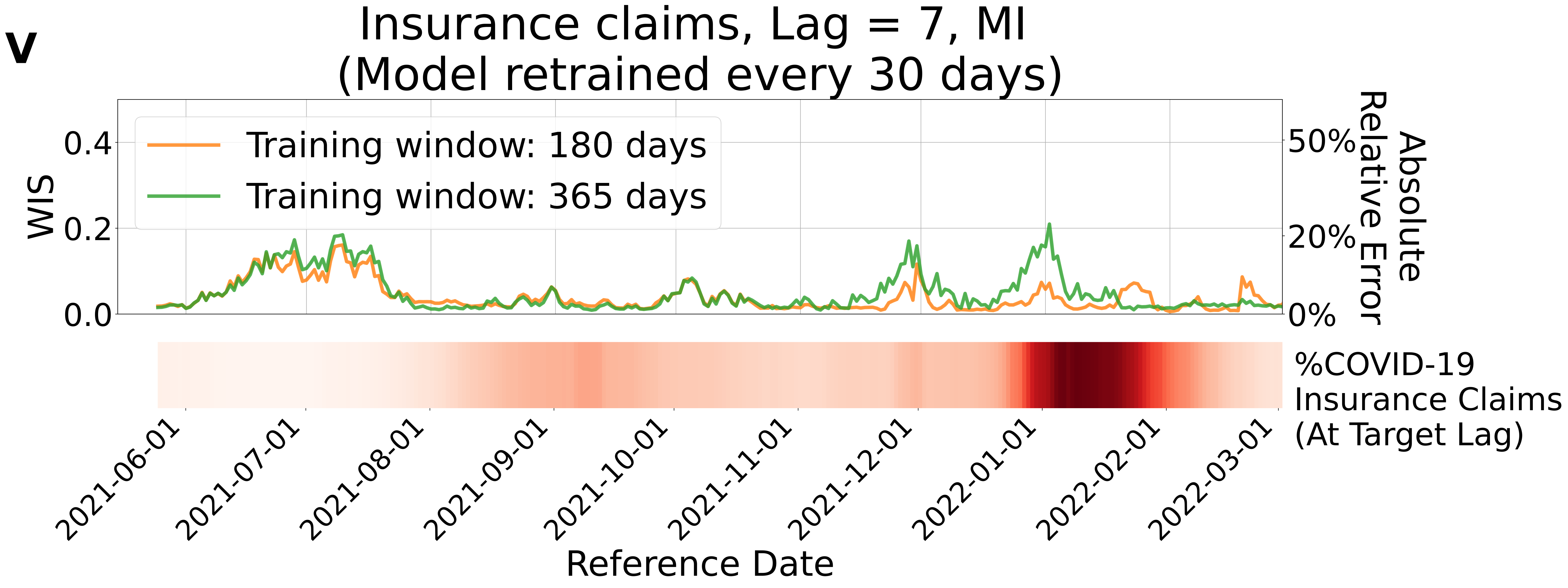
v
Insurance claims, Lag = 7, MI
(Model retrained every 30 days)
Training window: 180 days
Training window: 365 days
WIS
0.0
0.2
0.4
Absolute Relative Error
0%
20%
50%
%COVID-19 Insurance Claims (At Target Lag)
2021-06-01
2021-07-01
2021-08-01
2021-09-01
2021-10-01
2021-11-01
2021-12-01
2022-01-01
2022-02-01
2022-03-01
Reference Date

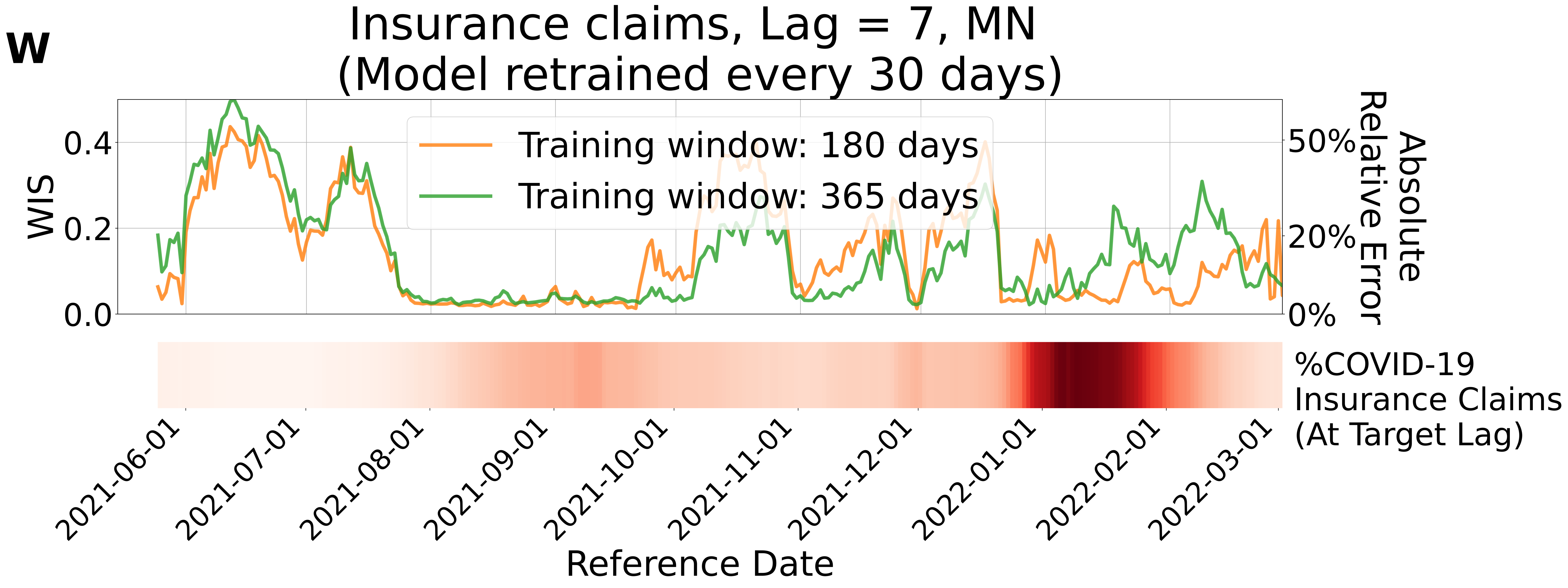

w
Insurance claims, Lag = 7, MN
(Model retrained every 30 days)
Training window: 180 days
Training window: 365 days
WIS
0.0
0.2
0.4
Absolute Relative Error
0%
20%
50%
%COVID-19 Insurance Claims (At Target Lag)
2021-06-01
2021-07-01
2021-08-01
2021-09-01
2021-10-01
2021-11-01
2021-12-01
2022-01-01
2022-02-01
2022-03-01
Reference Date

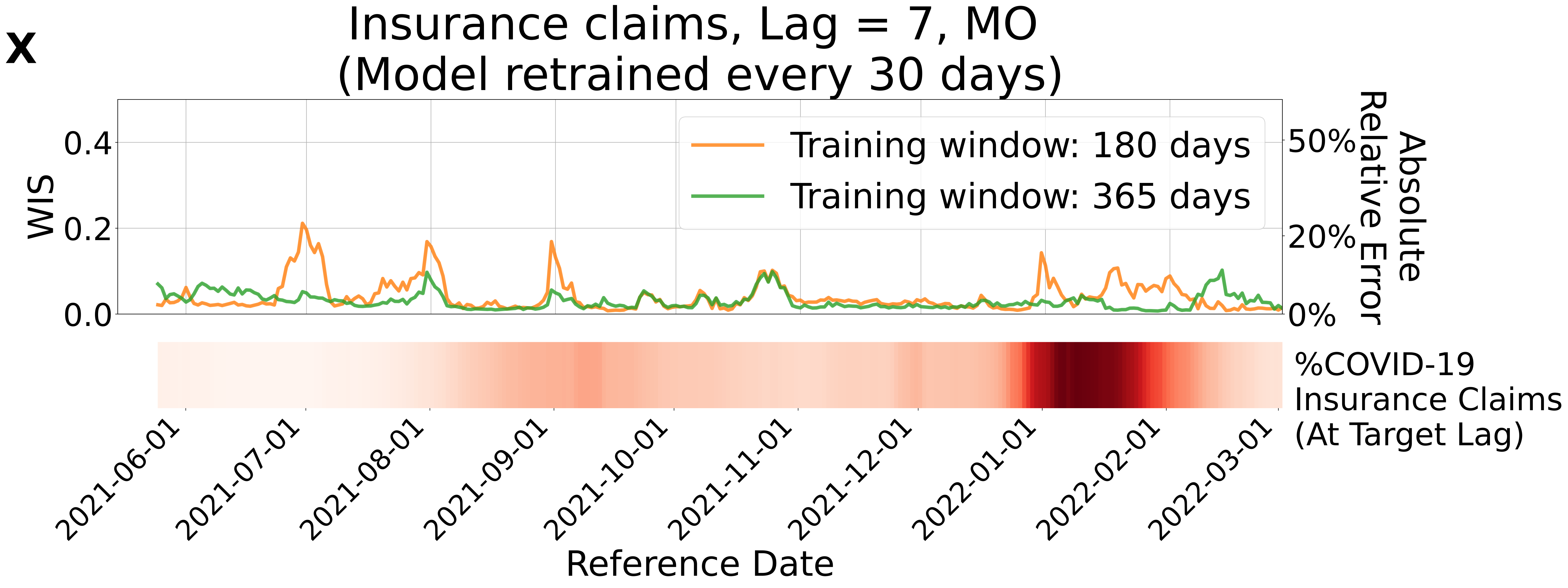
X
Insurance claims, Lag = 7, MO
(Model retrained every 30 days)
WIS
0.4
0.2
0.0
50%
20%
0%
Absolute Relative Error
Training window: 180 days
Training window: 365 days
%COVID-19 Insurance Claims (At Target Lag)
2021-06-01
2021-07-01
2021-08-01
2021-09-01
2021-10-01
2021-11-01
2021-12-01
2022-01-01
2022-02-01
2022-03-01
Reference Date

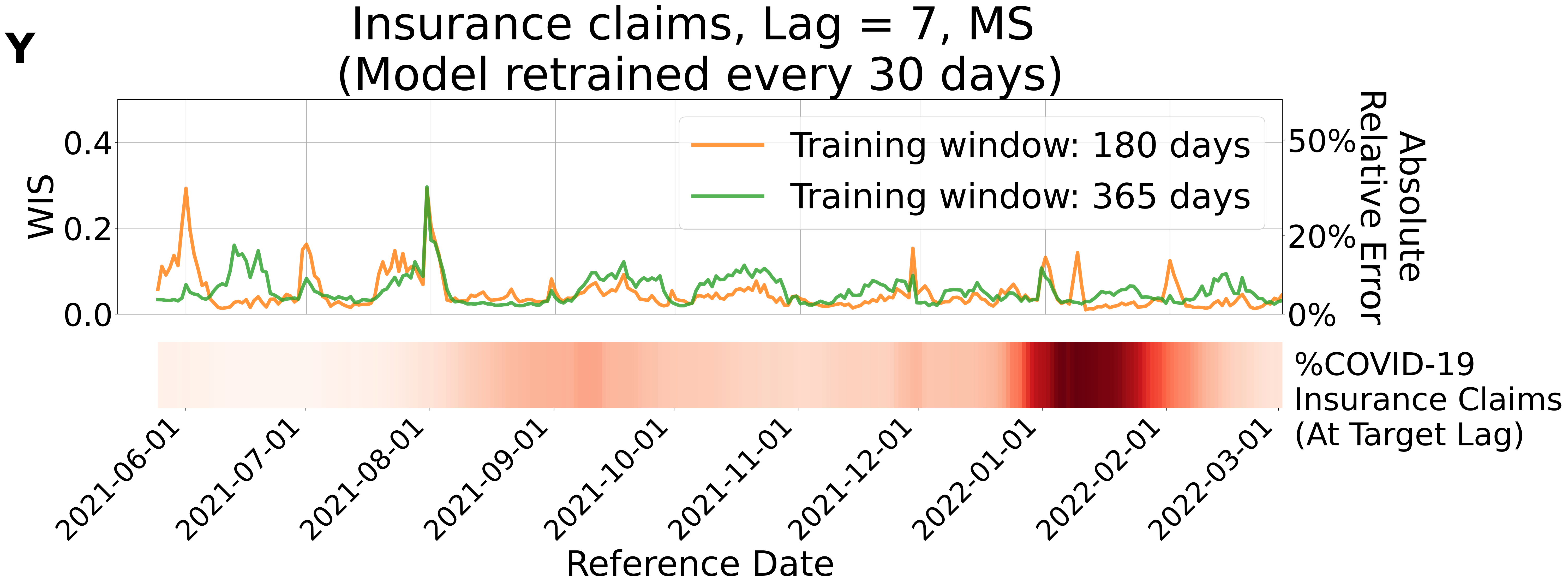

Y
Insurance claims, Lag = 7, MS
(Model retrained every 30 days)
WIS
0.0
0.2
0.4
Absolute Relative Error
0%
20%
50%
Training window: 180 days
Training window: 365 days
%COVID-19 Insurance Claims (At Target Lag)
2021-06-01
2021-07-01
2021-08-01
2021-09-01
2021-10-01
2021-11-01
2021-12-01
2022-01-01
2022-02-01
2022-03-01
Reference Date

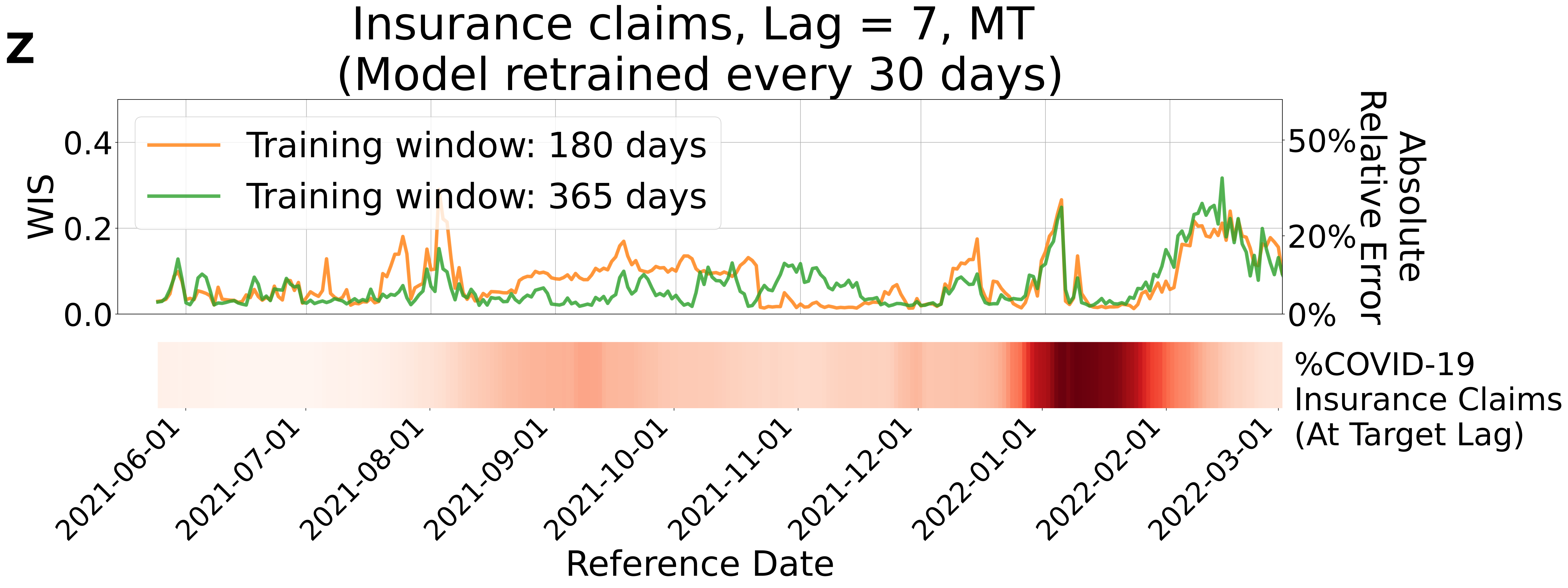
z
Insurance claims, Lag = 7, MT
(Model retrained every 30 days)
Training window: 180 days
Training window: 365 days
WIS
0.0
0.2
0.4
Absolute Relative Error
0%
20%
50%
%COVID-19
Insurance Claims
(At Target Lag)
2021-06-01
2021-07-01
2021-08-01
2021-09-01
2021-10-01
2021-11-01
2021-12-01
2022-01-01
2022-02-01
2022-03-01
Reference Date

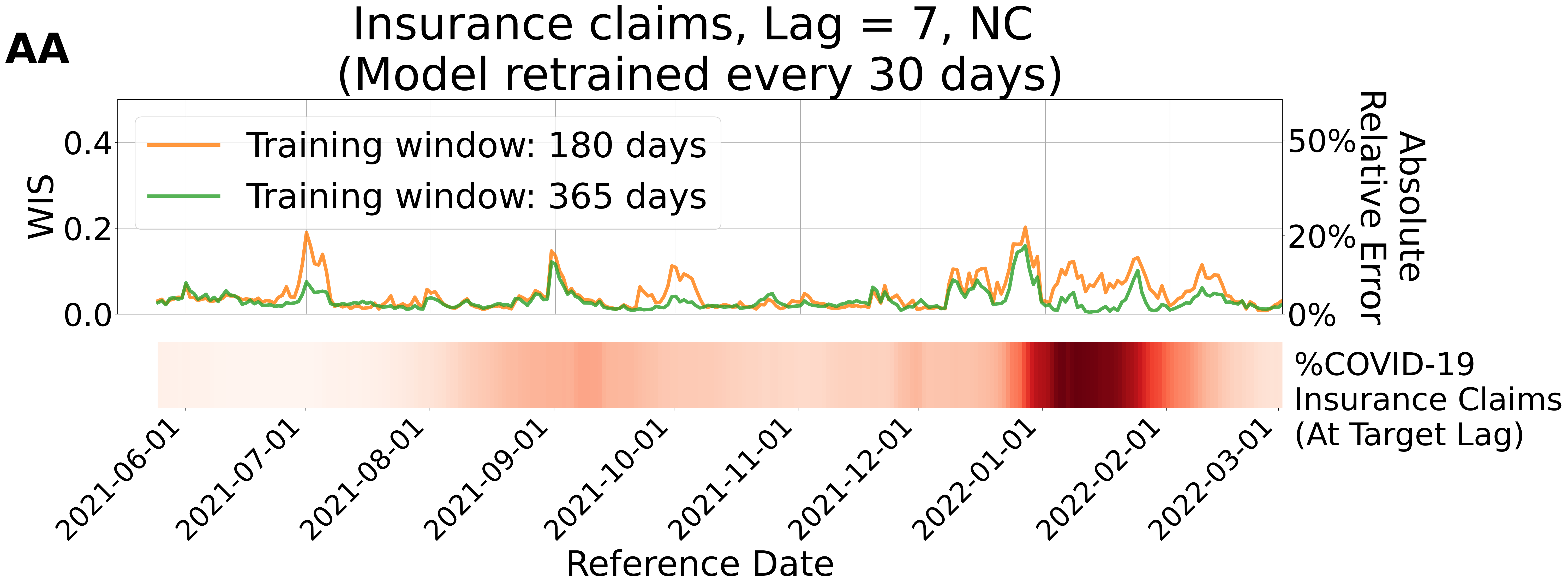
AA
Insurance claims, Lag = 7, NC
(Model retrained every 30 days)
Training window: 180 days
Training window: 365 days
WIS
0.4
0.2
0.0
Absolute Relative Error
50%
20%
0%
%COVID-19 Insurance Claims (At Target Lag)
2021-06-01
2021-07-01
2021-08-01
2021-09-01
2021-10-01
2021-11-01
2021-12-01
2022-01-01
2022-02-01
2022-03-01
Reference Date

**AB**

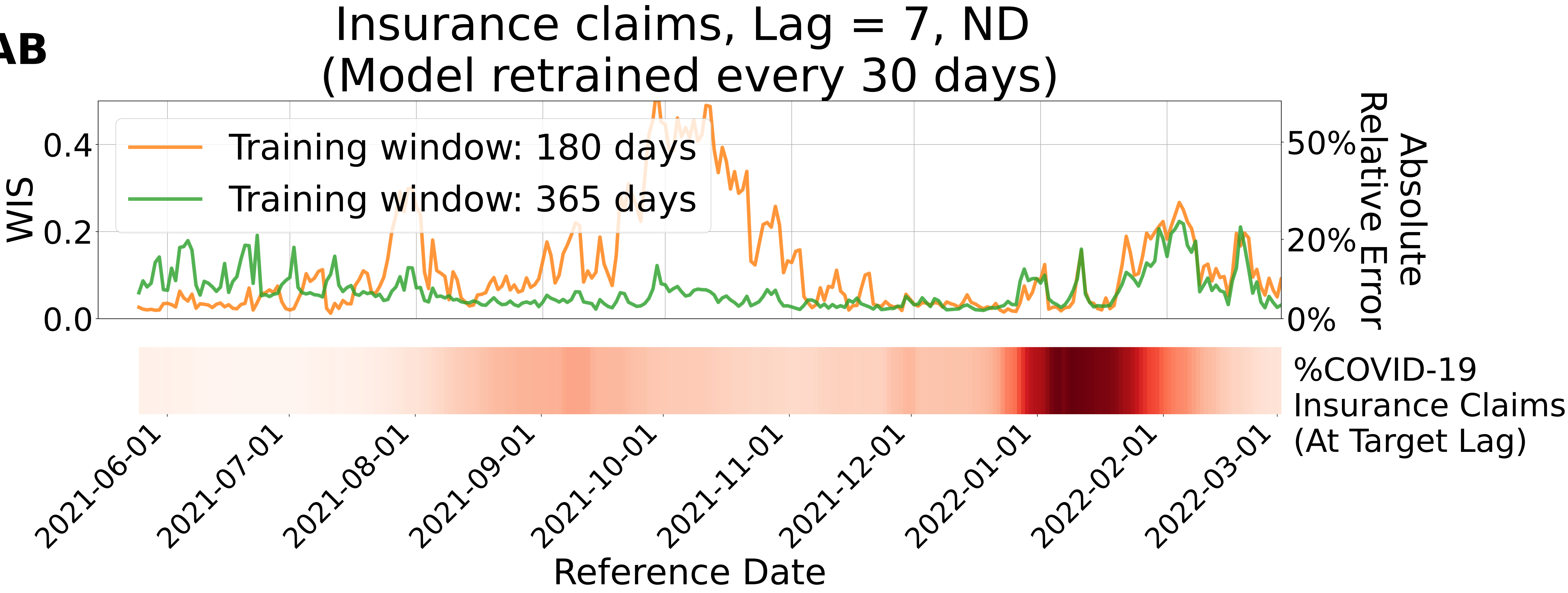

Insurance claims, Lag = 7, ND
(Model retrained every 30 days)
Training window: 180 days
Training window: 365 days
WIS
0.4
0.2
0.0
Absolute Relative Error
50%
20%
0%
%COVID-19 Insurance Claims (At Target Lag)
2021-06-01
2021-07-01
2021-08-01
2021-09-01
2021-10-01
2021-11-01
2021-12-01
2022-01-01
2022-02-01
2022-03-01
Reference Date

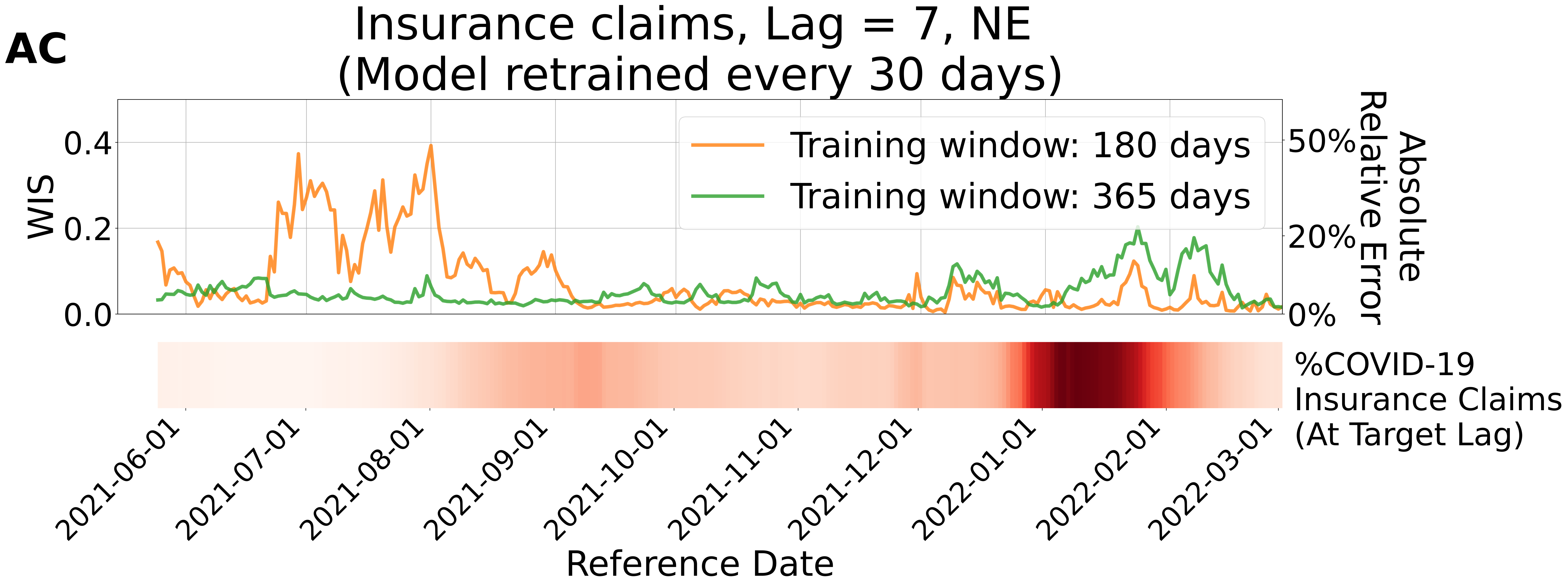

AC
Insurance claims, Lag = 7, NE
(Model retrained every 30 days)
WIS
0.0
0.2
0.4
Absolute Relative Error
0%
20%
50%
Training window: 180 days
Training window: 365 days
%COVID-19 Insurance Claims (At Target Lag)
2021-06-01
2021-07-01
2021-08-01
2021-09-01
2021-10-01
2021-11-01
2021-12-01
2022-01-01
2022-02-01
2022-03-01
Reference Date

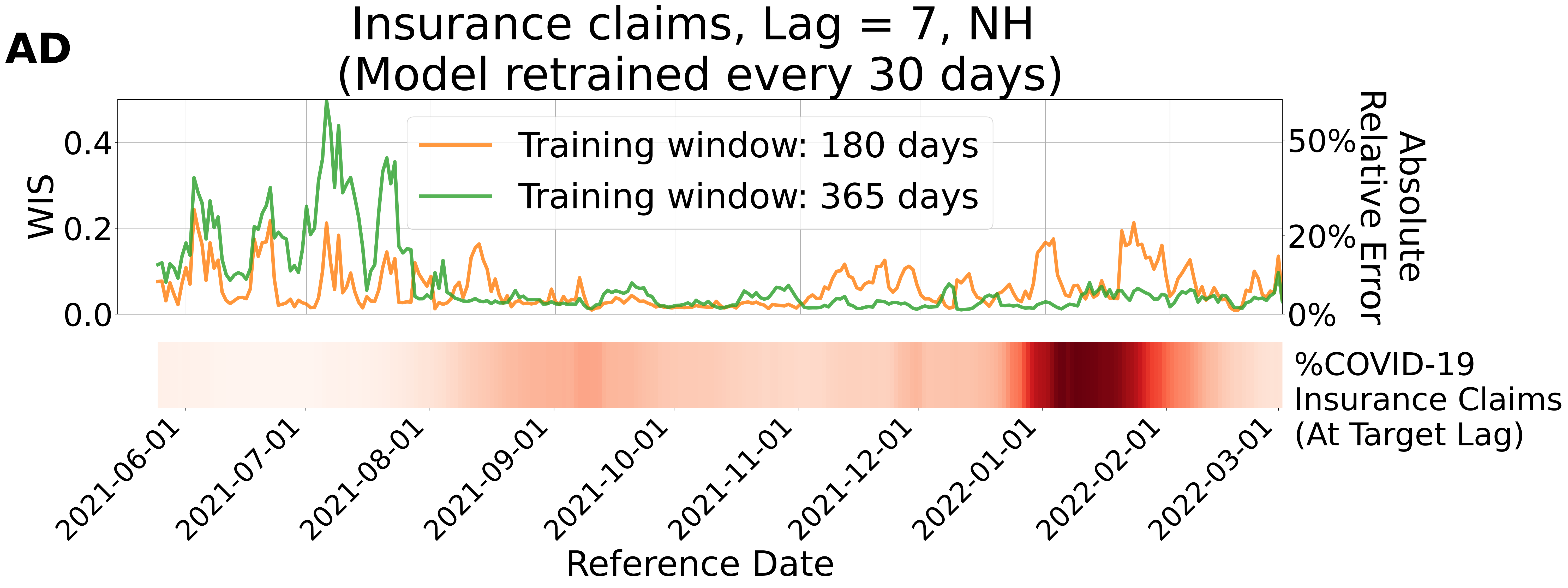
AD
Insurance claims, Lag = 7, NH
(Model retrained every 30 days)
WIS
0.4
0.2
0.0
Absolute Relative Error
50%
20%
0%
Training window: 180 days
Training window: 365 days
%COVID-19 Insurance Claims (At Target Lag)
2021-06-01
2021-07-01
2021-08-01
2021-09-01
2021-10-01
2021-11-01
2021-12-01
2022-01-01
2022-02-01
2022-03-01
Reference Date

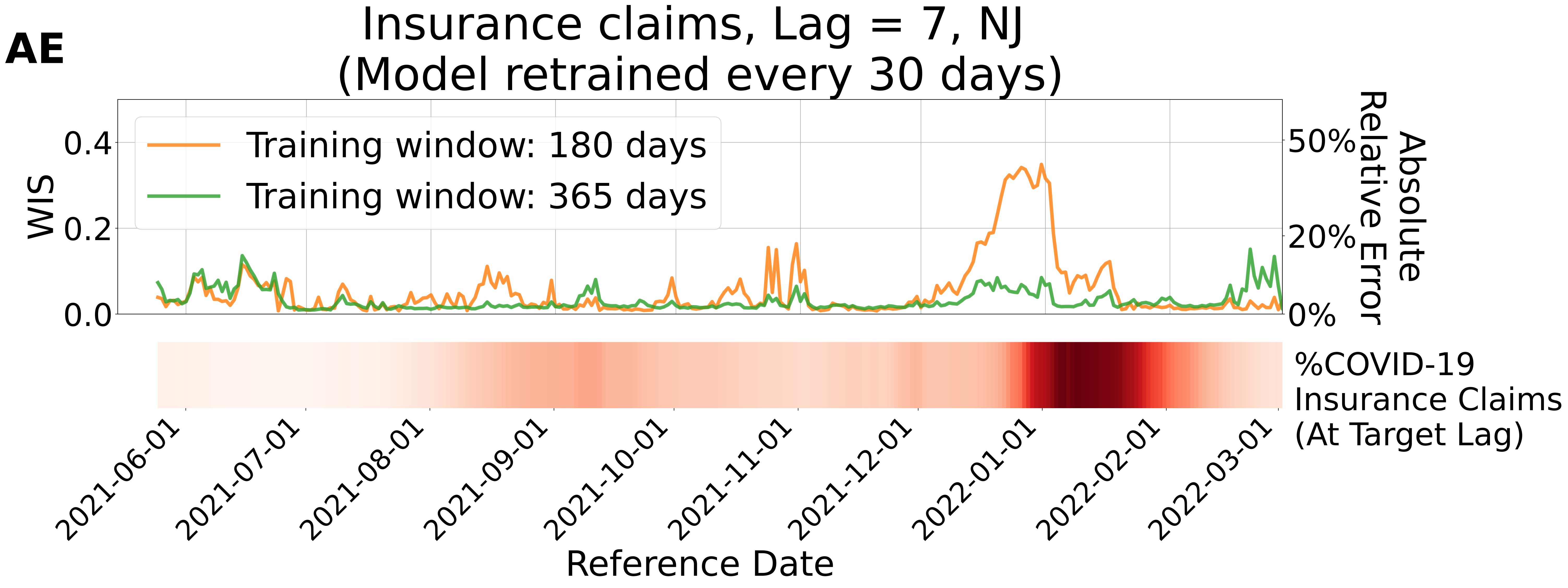

AE
Insurance claims, Lag = 7, NJ
(Model retrained every 30 days)
Training window: 180 days
Training window: 365 days
WIS
0.0
0.2
0.4
Absolute Relative Error
0%
20%
50%
%COVID-19 Insurance Claims (At Target Lag)
2021-06-01
2021-07-01
2021-08-01
2021-09-01
2021-10-01
2021-11-01
2021-12-01
2022-01-01
2022-02-01
2022-03-01
Reference Date

**AF**

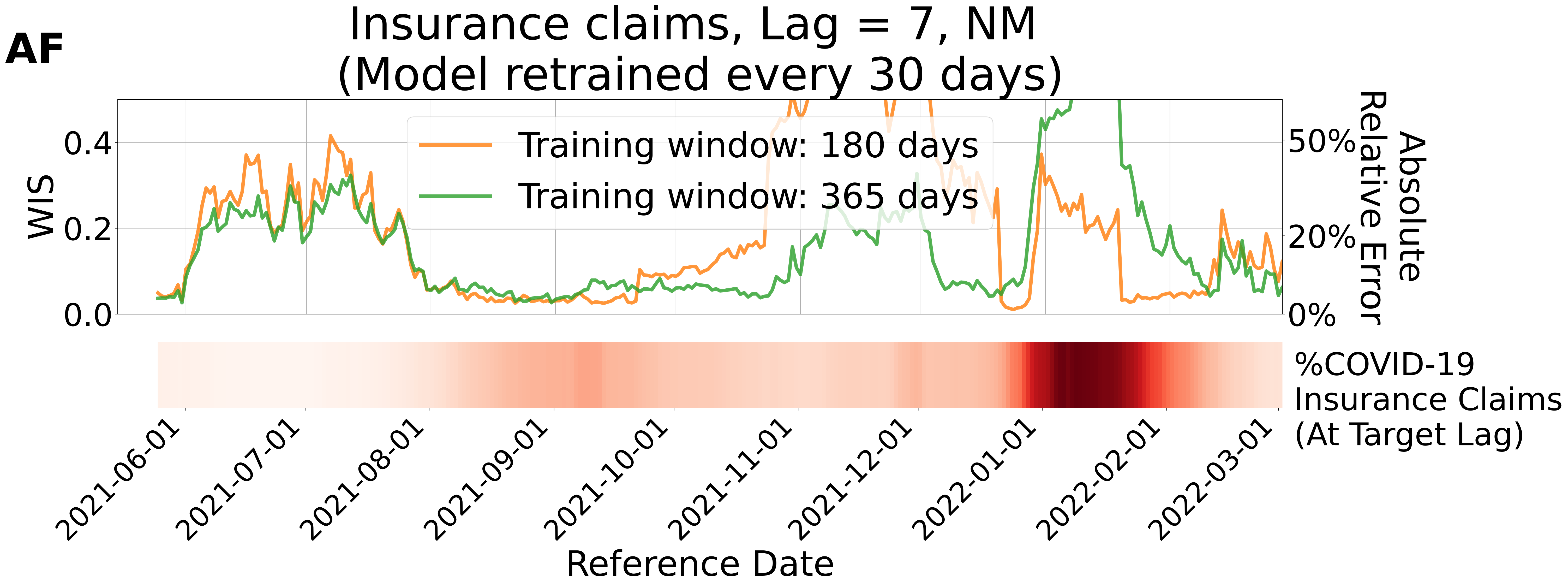
Insurance claims, Lag = 7, NM
(Model retrained every 30 days)
Training window: 180 days
Training window: 365 days
WIS
0.0
0.2
0.4
Absolute Relative Error
0%
20%
50%
%COVID-19 Insurance Claims (At Target Lag)
2021-06-01
2021-07-01
2021-08-01
2021-09-01
2021-10-01
2021-11-01
2021-12-01
2022-01-01
2022-02-01
2022-03-01
Reference Date

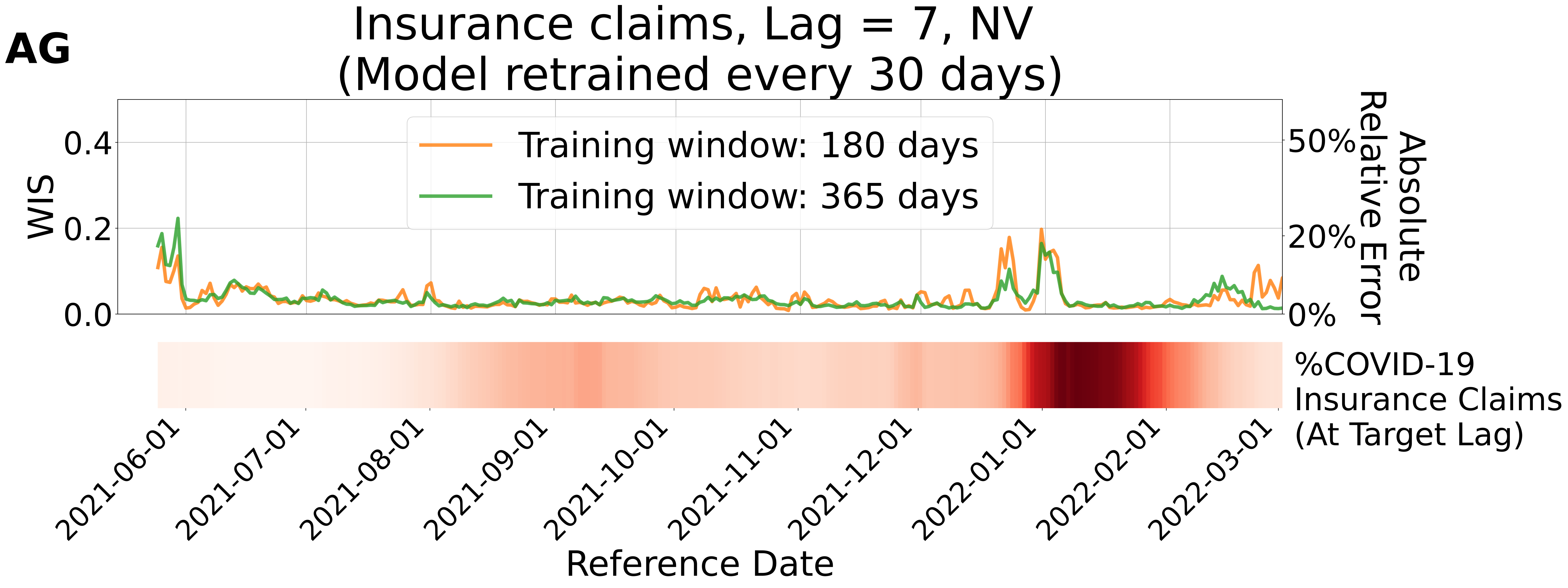
AG
Insurance claims, Lag = 7, NV
(Model retrained every 30 days)
Training window: 180 days
Training window: 365 days
WIS
0.0
0.2
0.4
Absolute Relative Error
0%
20%
50%
%COVID-19 Insurance Claims (At Target Lag)
2021-06-01
2021-07-01
2021-08-01
2021-09-01
2021-10-01
2021-11-01
2021-12-01
2022-01-01
2022-02-01
2022-03-01
Reference Date

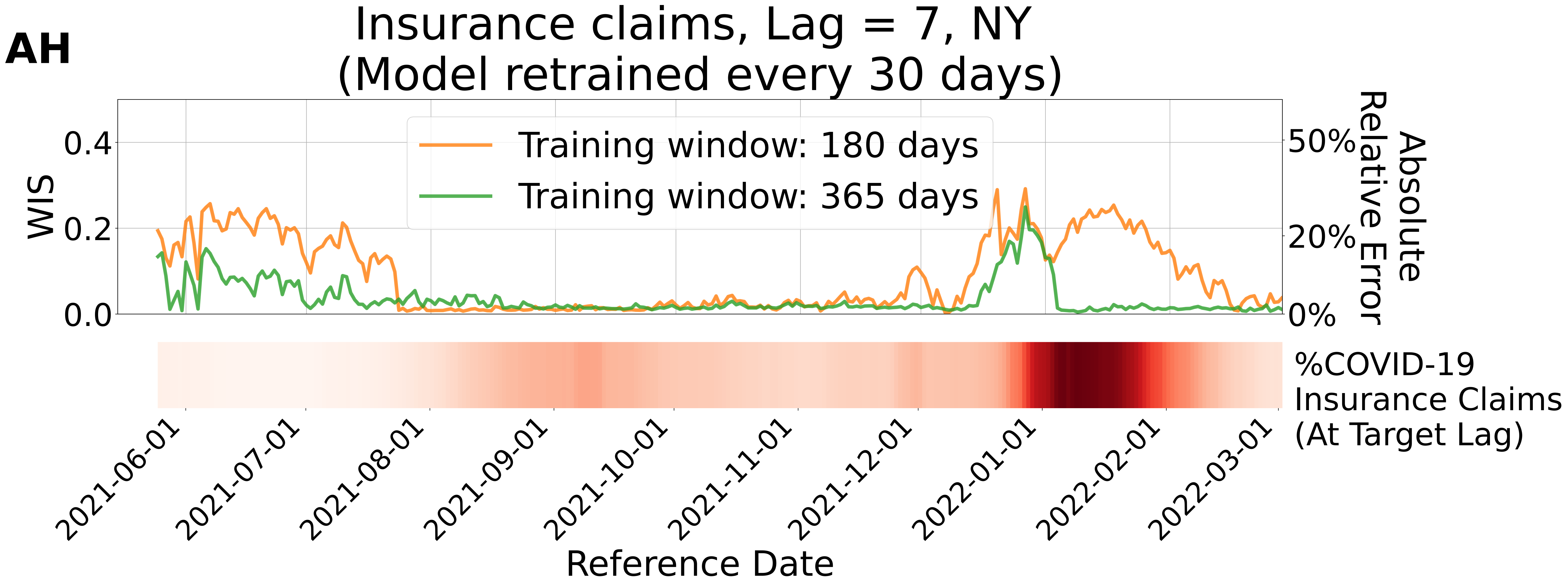
AH
Insurance claims, Lag = 7, NY
(Model retrained every 30 days)
Training window: 180 days
Training window: 365 days
WIS
0.0
0.2
0.4
Absolute Relative Error
0%
20%
50%
%COVID-19 Insurance Claims (At Target Lag)
2021-06-01
2021-07-01
2021-08-01
2021-09-01
2021-10-01
2021-11-01
2021-12-01
2022-01-01
2022-02-01
2022-03-01
Reference Date

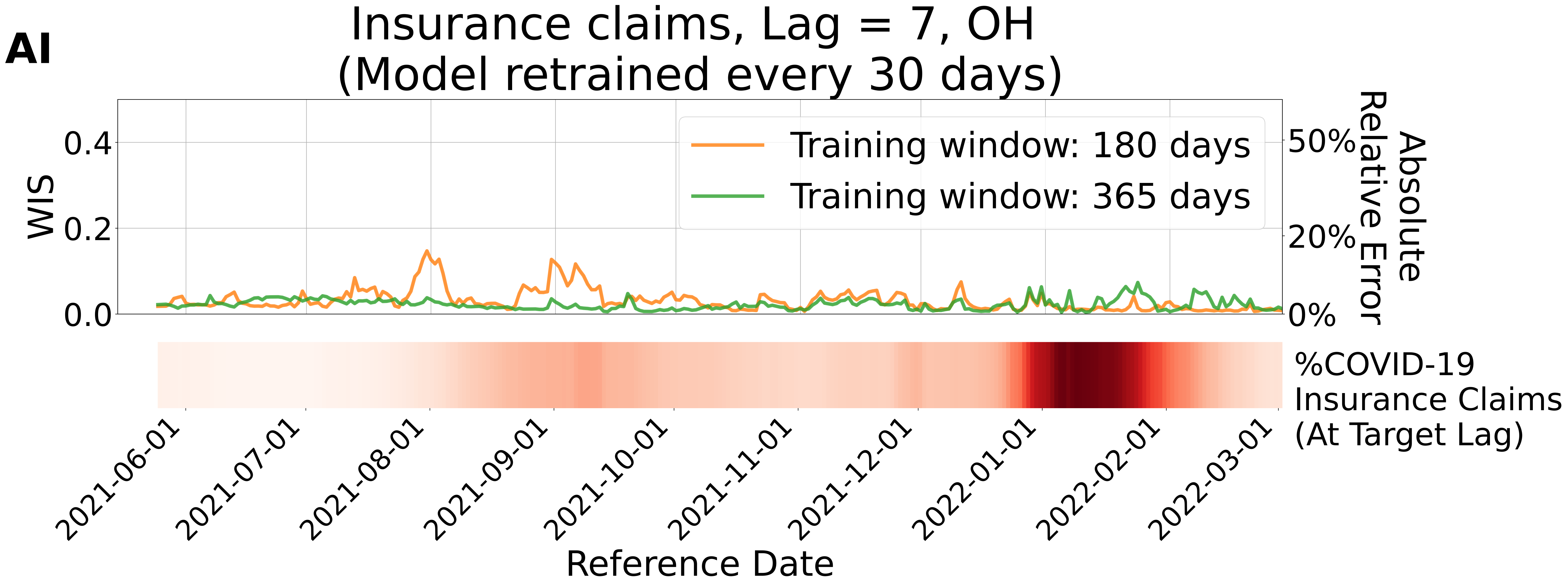
AI
Insurance claims, Lag = 7, OH
(Model retrained every 30 days)
WIS
0.0
0.2
0.4
Absolute Relative Error
0%
20%
50%
Training window: 180 days
Training window: 365 days
%COVID-19 Insurance Claims (At Target Lag)
2021-06-01
2021-07-01
2021-08-01
2021-09-01
2021-10-01
2021-11-01
2021-12-01
2022-01-01
2022-02-01
2022-03-01
Reference Date

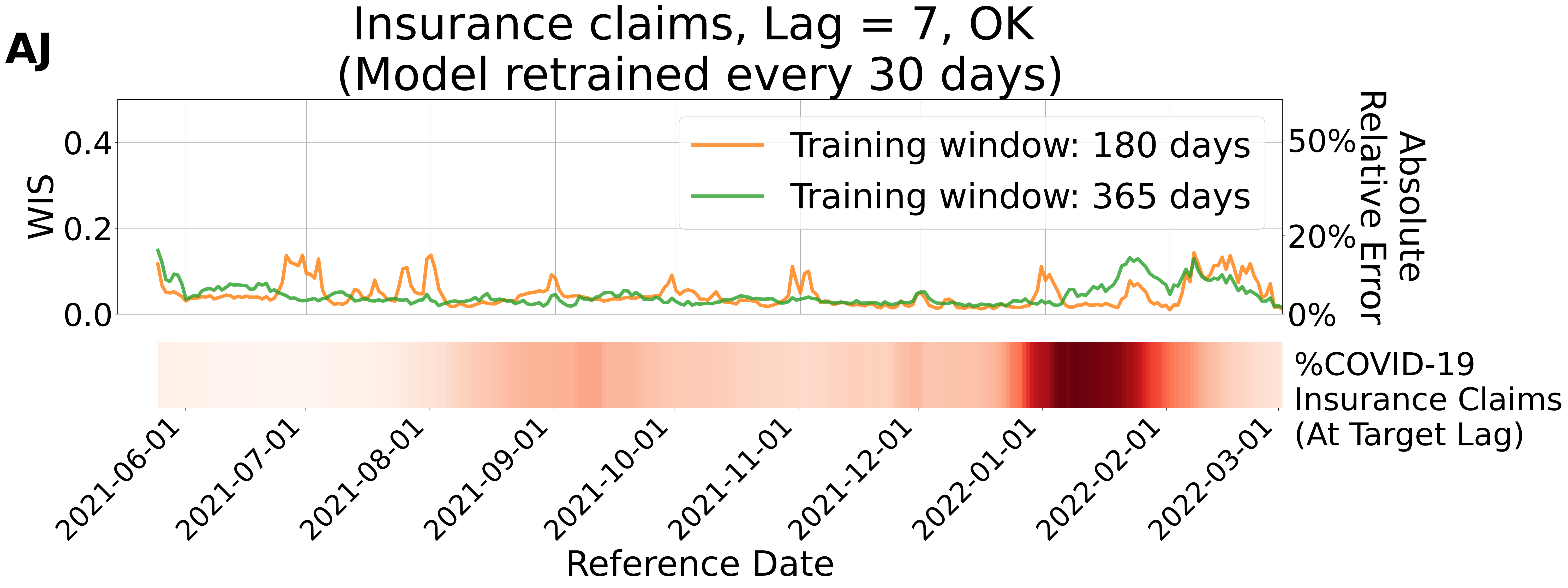
AJ
Insurance claims, Lag = 7, OK
(Model retrained every 30 days)
WIS
0.0
0.2
0.4
Absolute Relative Error
0%
20%
50%
Training window: 180 days
Training window: 365 days
%COVID-19 Insurance Claims (At Target Lag)
2021-06-01
2021-07-01
2021-08-01
2021-09-01
2021-10-01
2021-11-01
2021-12-01
2022-01-01
2022-02-01
2022-03-01
Reference Date

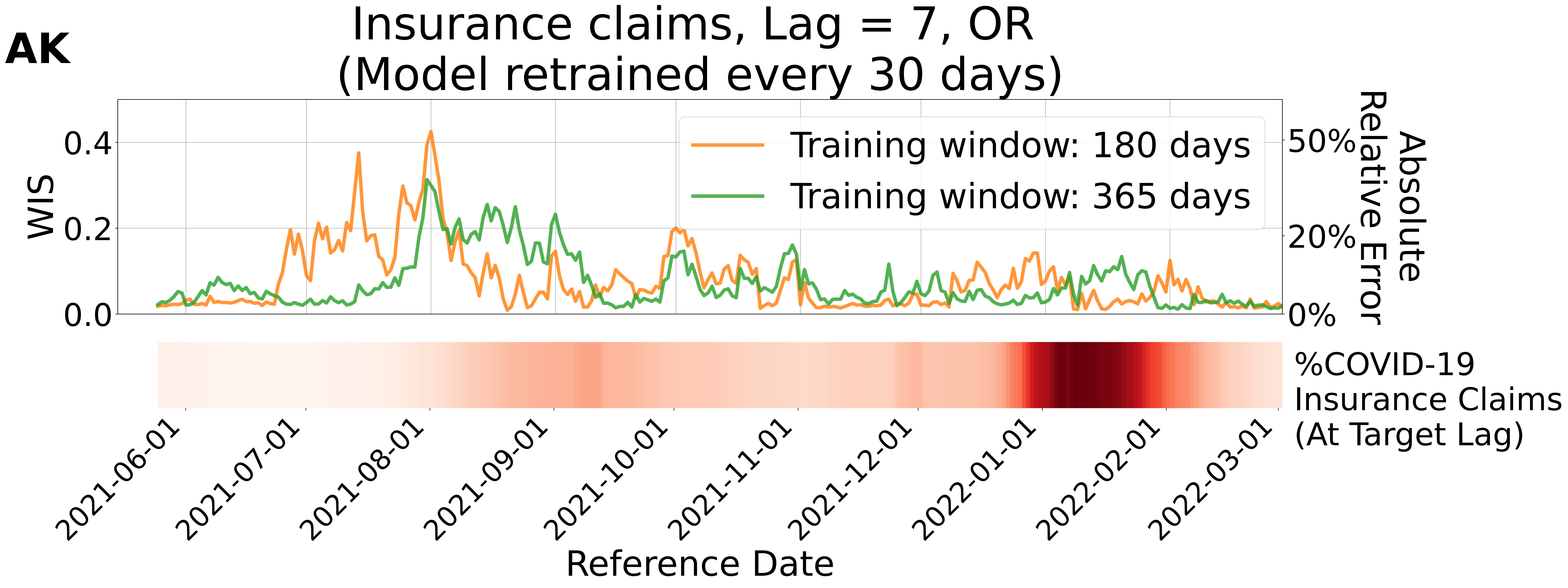

AK
Insurance claims, Lag = 7, OR
(Model retrained every 30 days)
WIS
0.0
0.2
0.4
Absolute Relative Error
0%
20%
50%
Training window: 180 days
Training window: 365 days
%COVID-19 Insurance Claims (At Target Lag)
2021-06-01
2021-07-01
2021-08-01
2021-09-01
2021-10-01
2021-11-01
2021-12-01
2022-01-01
2022-02-01
2022-03-01
Reference Date

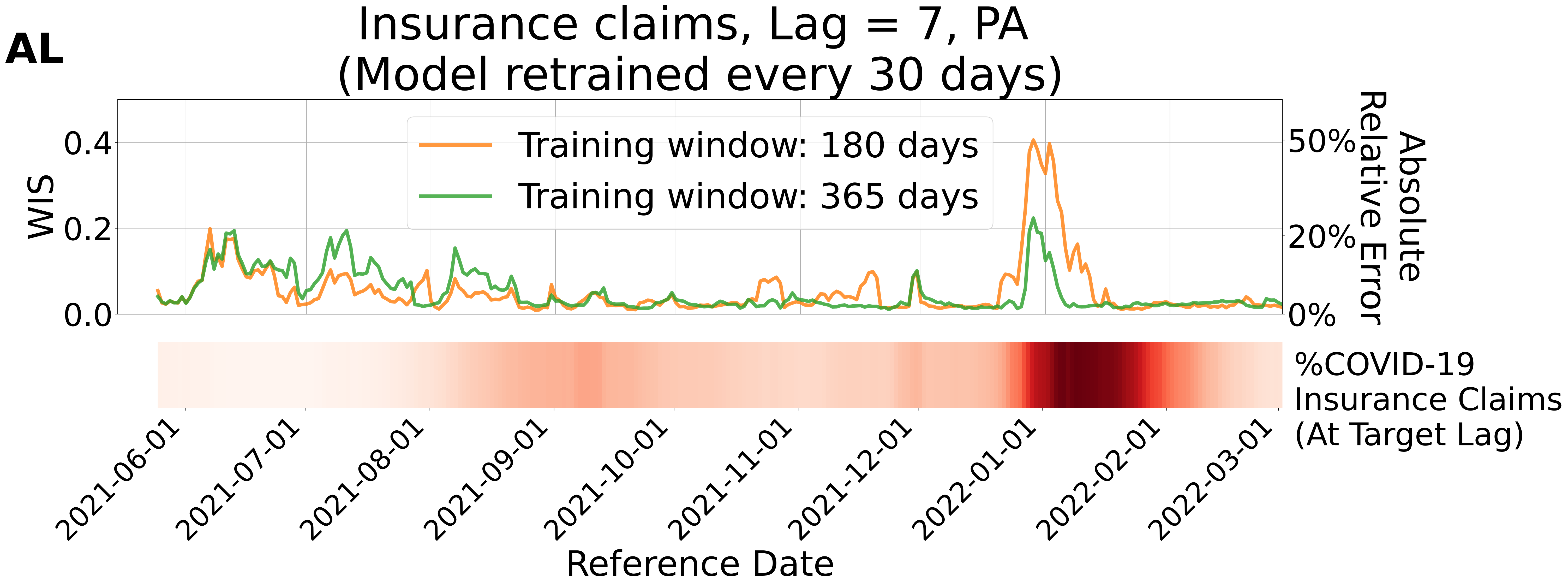
AL
Insurance claims, Lag = 7, PA
(Model retrained every 30 days)
Training window: 180 days
Training window: 365 days
WIS
0.0
0.2
0.4
Absolute Relative Error
0%
20%
50%
%COVID-19 Insurance Claims (At Target Lag)
2021-06-01
2021-07-01
2021-08-01
2021-09-01
2021-10-01
2021-11-01
2021-12-01
2022-01-01
2022-02-01
2022-03-01
Reference Date

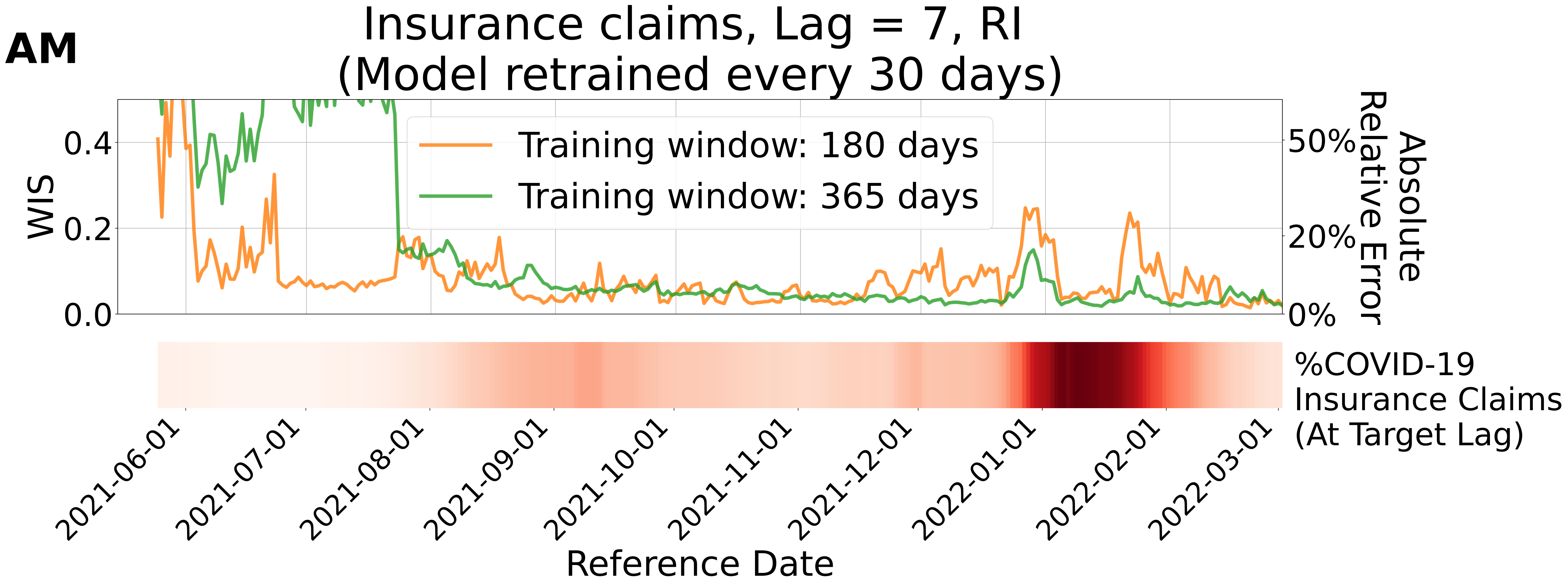

AM
Insurance claims, Lag = 7, RI
(Model retrained every 30 days)
Training window: 180 days
Training window: 365 days
WIS
0.4
0.2
0.0
Absolute Relative Error
50%
20%
0%
%COVID-19 Insurance Claims (At Target Lag)
2021-06-01
2021-07-01
2021-08-01
2021-09-01
2021-10-01
2021-11-01
2021-12-01
2022-01-01
2022-02-01
2022-03-01
Reference Date

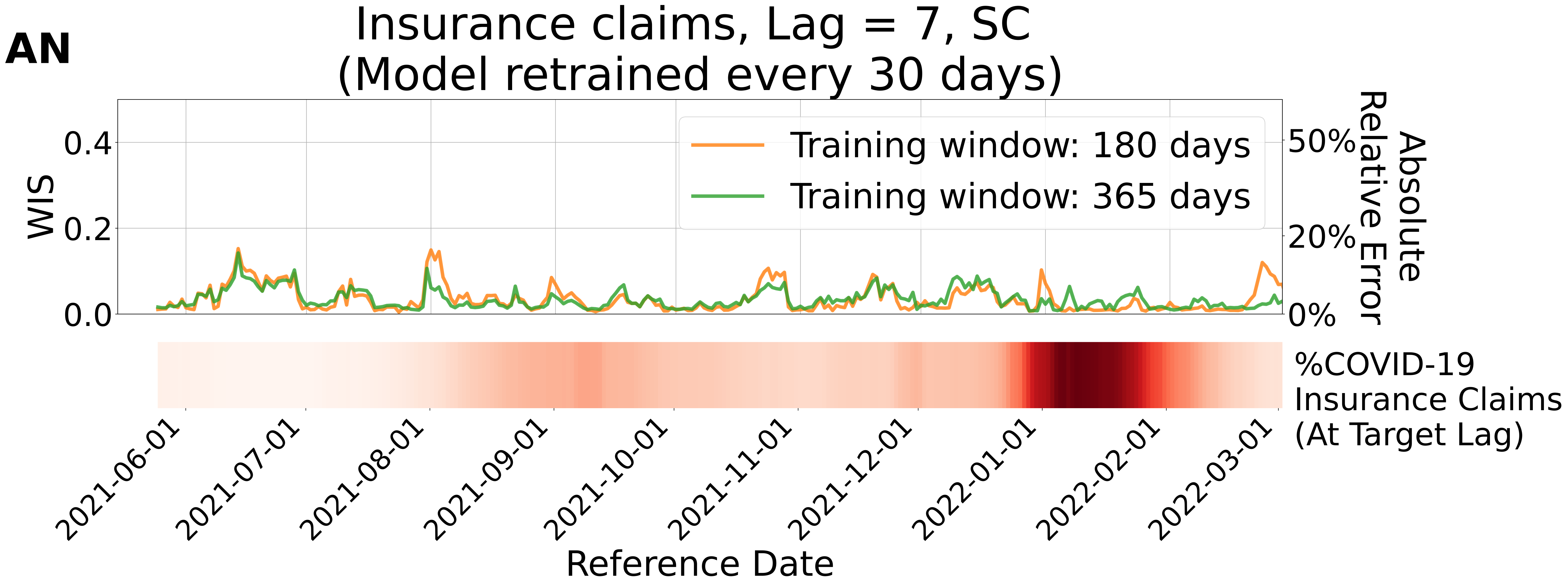
AN
Insurance claims, Lag = 7, SC
(Model retrained every 30 days)
WIS
0.4
0.2
0.0
Absolute
Relative Error
50%
20%
0%
Training window: 180 days
Training window: 365 days
%COVID-19
Insurance Claims
(At Target Lag)
2021-06-01
2021-07-01
2021-08-01
2021-09-01
2021-10-01
2021-11-01
2021-12-01
2022-01-01
2022-02-01
2022-03-01
Reference Date

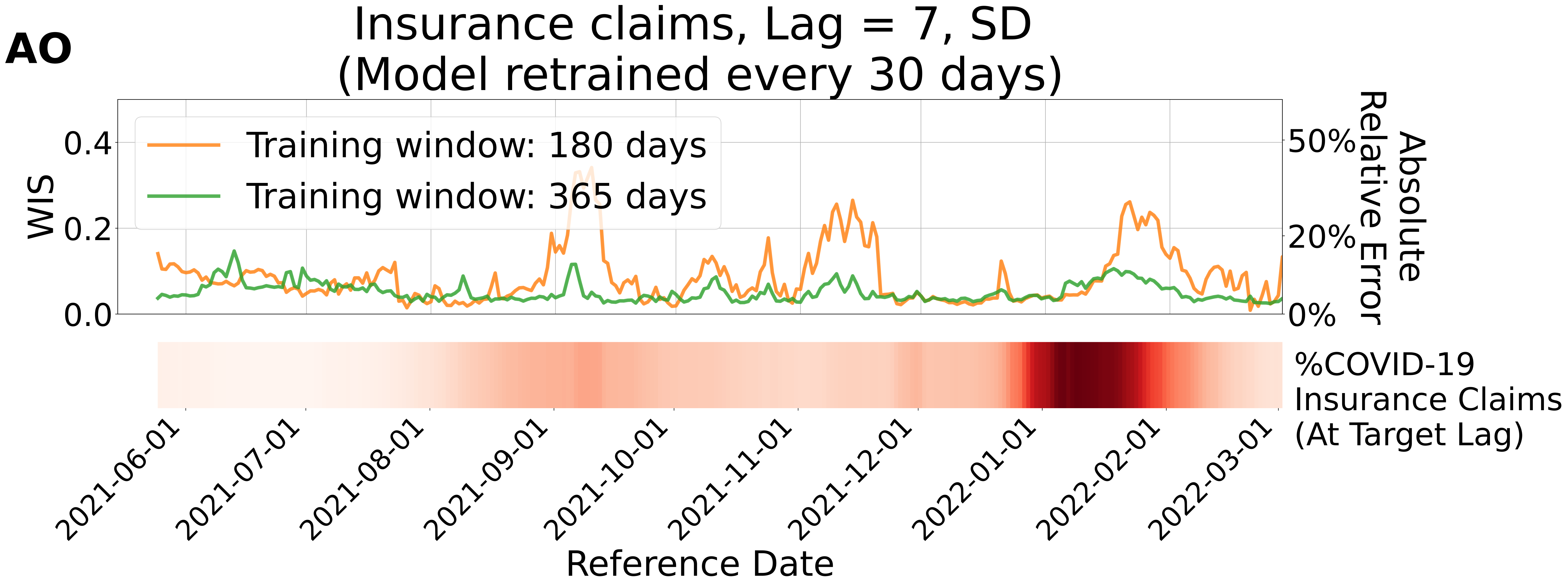
AO
Insurance claims, Lag = 7, SD
(Model retrained every 30 days)
Training window: 180 days
Training window: 365 days
WIS
0.0
0.2
0.4
0%
20%
50%
Absolute
Relative Error
%COVID-19
Insurance Claims
(At Target Lag)
2021-06-01
2021-07-01
2021-08-01
2021-09-01
2021-10-01
2021-11-01
2021-12-01
2022-01-01
2022-02-01
2022-03-01
Reference Date

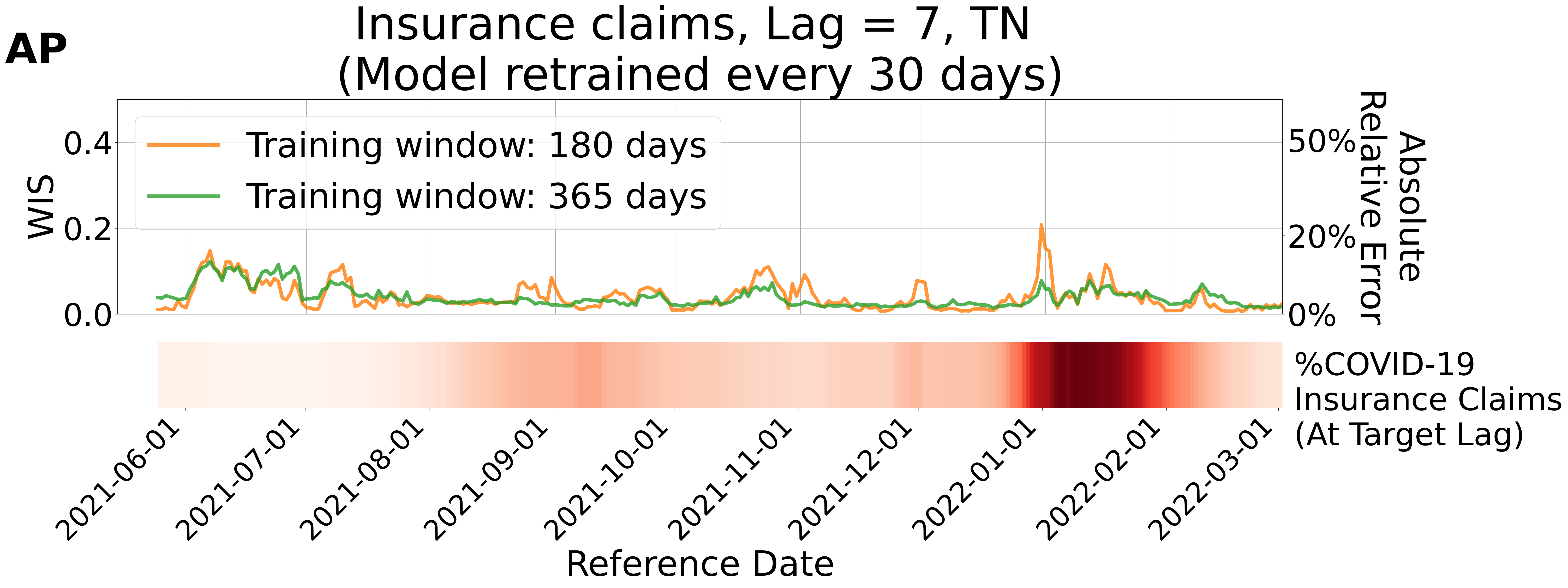
AP
Insurance claims, Lag = 7, TN
(Model retrained every 30 days)
Training window: 180 days
Training window: 365 days
WIS
0.4
0.2
0.0
Absolute Relative Error
50%
20%
0%
%COVID-19 Insurance Claims (At Target Lag)
2021-06-01
2021-07-01
2021-08-01
2021-09-01
2021-10-01
2021-11-01
2021-12-01
2022-01-01
2022-02-01
2022-03-01
Reference Date

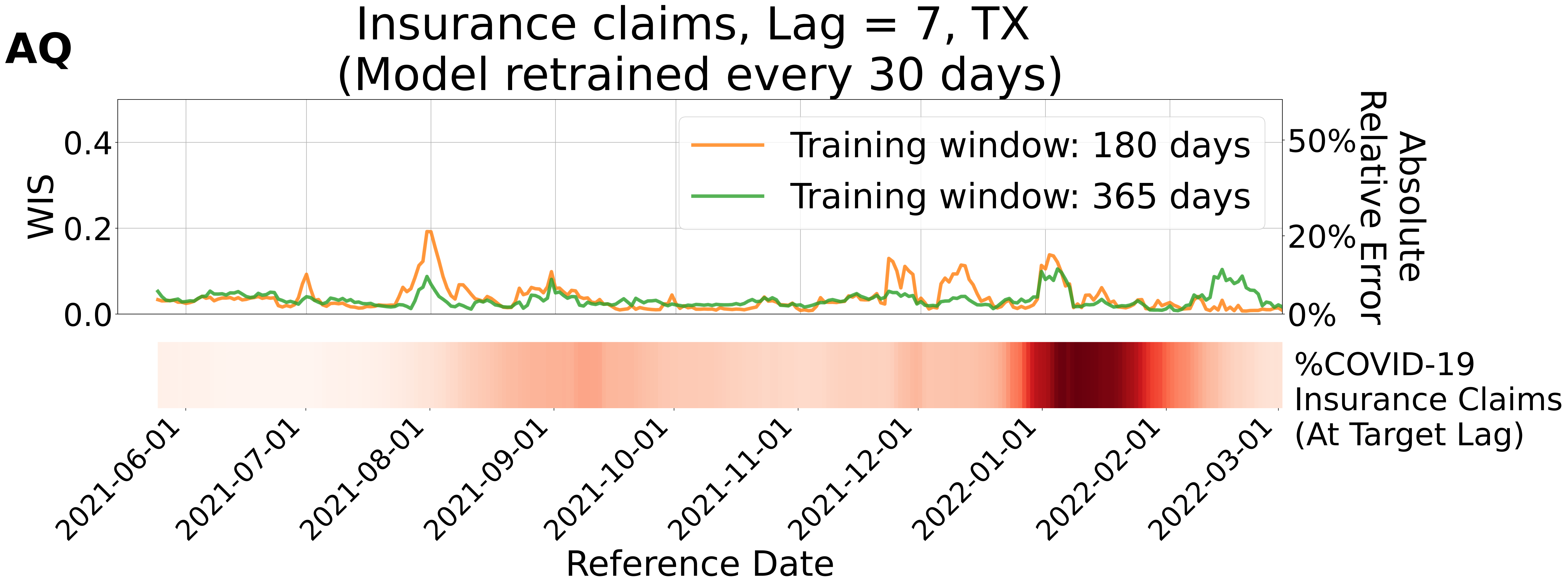

AQ
Insurance claims, Lag = 7, TX
(Model retrained every 30 days)
WIS
0.4
0.2
0.0
Absolute Relative Error
50%
20%
0%
Training window: 180 days
Training window: 365 days
%COVID-19
Insurance Claims
(At Target Lag)
2021-06-01
2021-07-01
2021-08-01
2021-09-01
2021-10-01
2021-11-01
2021-12-01
2022-01-01
2022-02-01
2022-03-01
Reference Date

**AR**

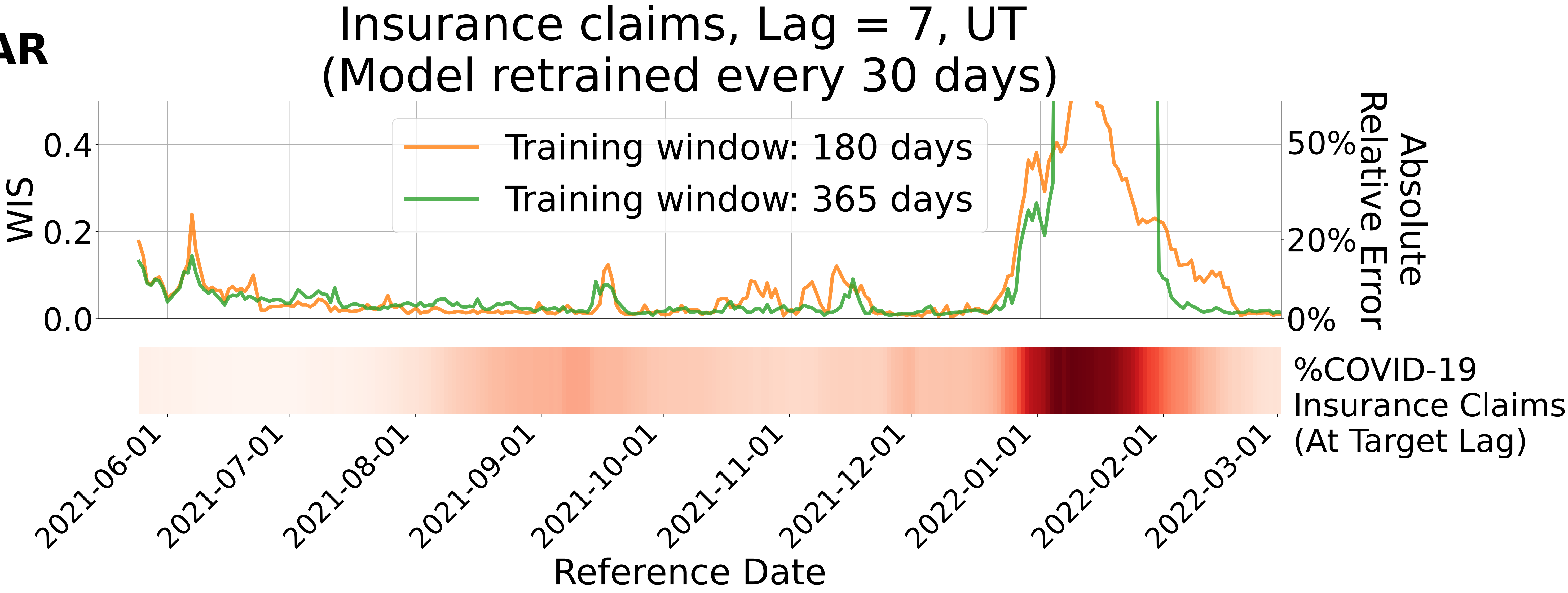

Insurance claims, Lag = 7, UT
(Model retrained every 30 days)
Training window: 180 days
Training window: 365 days
WIS
0.0
0.2
0.4
Absolute Relative Error
0%
20%
50%
%COVID-19 Insurance Claims (At Target Lag)
2021-06-01
2021-07-01
2021-08-01
2021-09-01
2021-10-01
2021-11-01
2021-12-01
2022-01-01
2022-02-01
2022-03-01
Reference Date

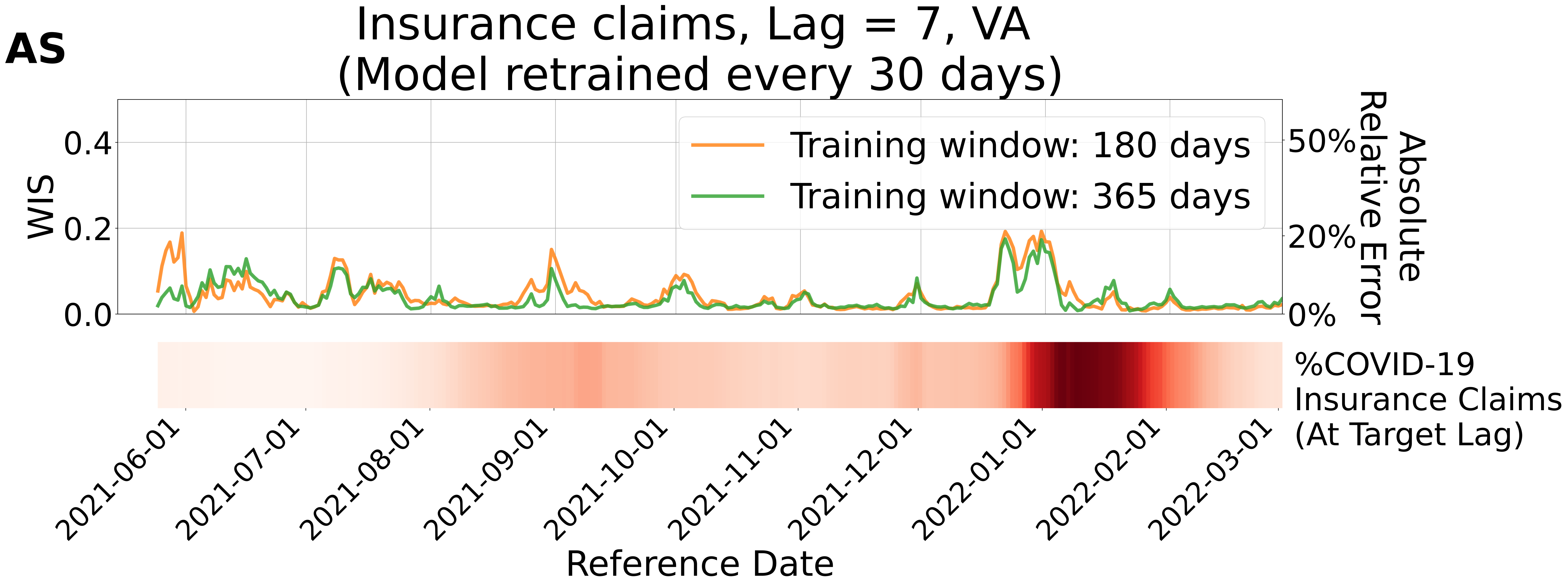
AS
Insurance claims, Lag = 7, VA
(Model retrained every 30 days)
WIS
0.0
0.2
0.4
Absolute Relative Error
0%
20%
50%
Training window: 180 days
Training window: 365 days
%COVID-19 Insurance Claims (At Target Lag)
2021-06-01
2021-07-01
2021-08-01
2021-09-01
2021-10-01
2021-11-01
2021-12-01
2022-01-01
2022-02-01
2022-03-01
Reference Date

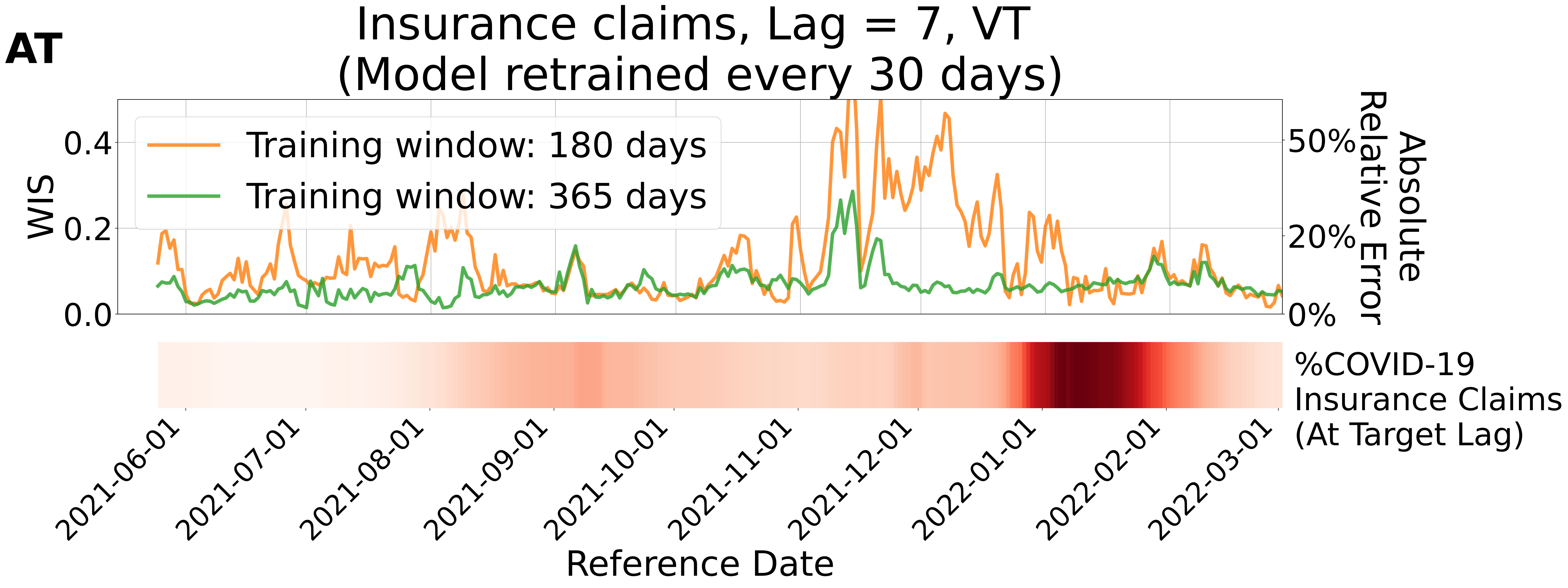
AT
Insurance claims, Lag = 7, VT
(Model retrained every 30 days)
Training window: 180 days
Training window: 365 days
WIS
0.0
0.2
0.4
Absolute Relative Error
0%
20%
50%
%COVID-19 Insurance Claims (At Target Lag)
2021-06-01
2021-07-01
2021-08-01
2021-09-01
2021-10-01
2021-11-01
2021-12-01
2022-01-01
2022-02-01
2022-03-01
Reference Date

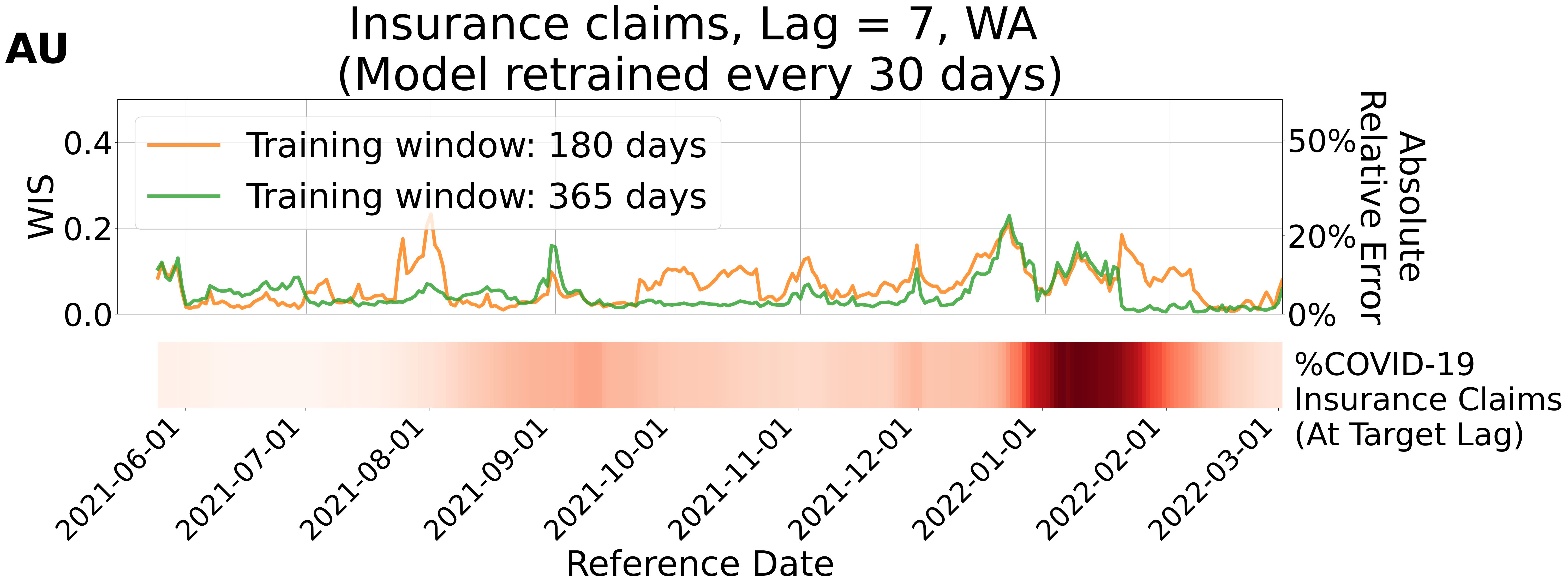

AU
Insurance claims, Lag = 7, WA
(Model retrained every 30 days)
Training window: 180 days
Training window: 365 days
WIS
0.4
0.2
0.0
Absolute Relative Error
50%
20%
0%
%COVID-19 Insurance Claims (At Target Lag)
2021-06-01
2021-07-01
2021-08-01
2021-09-01
2021-10-01
2021-11-01
2021-12-01
2022-01-01
2022-02-01
2022-03-01
Reference Date

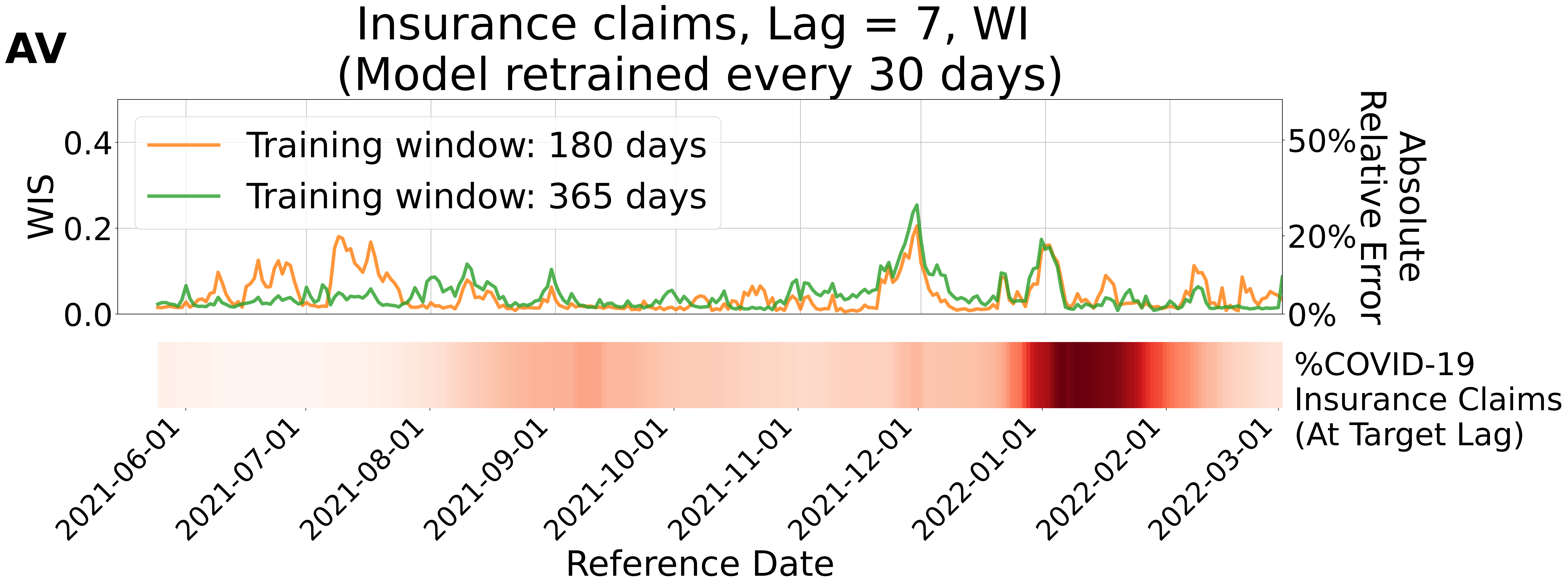
AV
Insurance claims, Lag = 7, WI
(Model retrained every 30 days)
Training window: 180 days
Training window: 365 days
WIS
0.0
0.2
0.4
Absolute Relative Error
0%
20%
50%
%COVID-19
Insurance Claims
(At Target Lag)
2021-06-01
2021-07-01
2021-08-01
2021-09-01
2021-10-01
2021-11-01
2021-12-01
2022-01-01
2022-02-01
2022-03-01
Reference Date

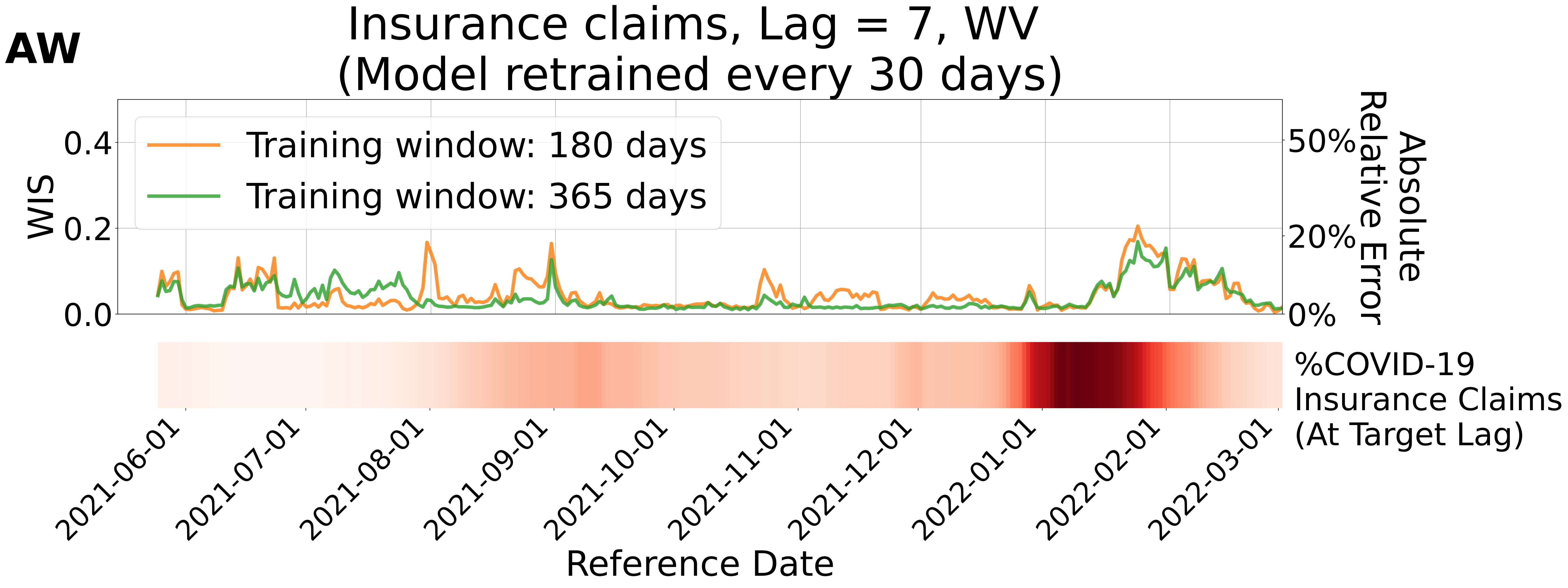
AW
Insurance claims, Lag = 7, WV
(Model retrained every 30 days)
Training window: 180 days
Training window: 365 days
WIS
0.0
0.2
0.4
Absolute
Relative Error
0%
20%
50%
%COVID-19
Insurance Claims
(At Target Lag)
2021-06-01
2021-07-01
2021-08-01
2021-09-01
2021-10-01
2021-11-01
2021-12-01
2022-01-01
2022-02-01
2022-03-01
Reference Date

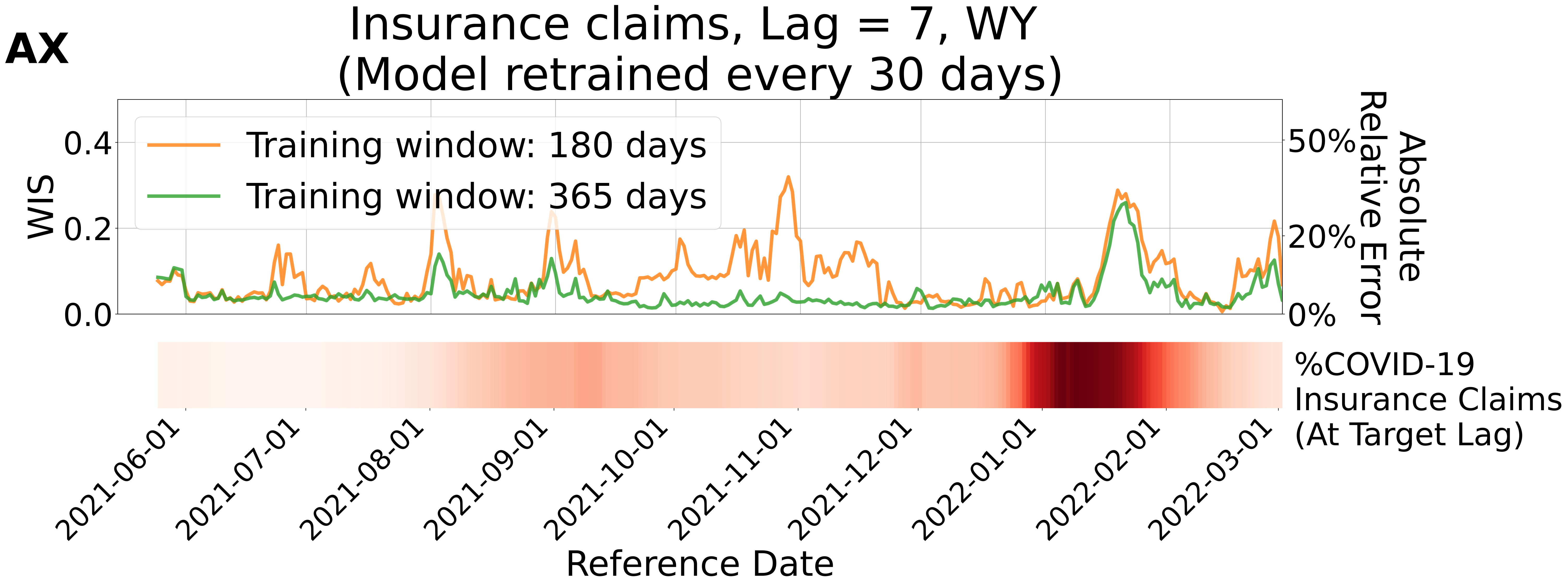
AX
Insurance claims, Lag = 7, WY
(Model retrained every 30 days)
Training window: 180 days
Training window: 365 days
WIS
0.0
0.2
0.4
0%
20%
50%
Absolute Relative Error
%COVID-19 Insurance Claims (At Target Lag)
2021-06-01
2021-07-01
2021-08-01
2021-09-01
2021-10-01
2021-11-01
2021-12-01
2022-01-01
2022-02-01
2022-03-01
Reference Date
