## Supplementary figures and images for "Real-time Forecasting of Data Revisions in Epidemic Surveillance Streams"

### S1_Fig

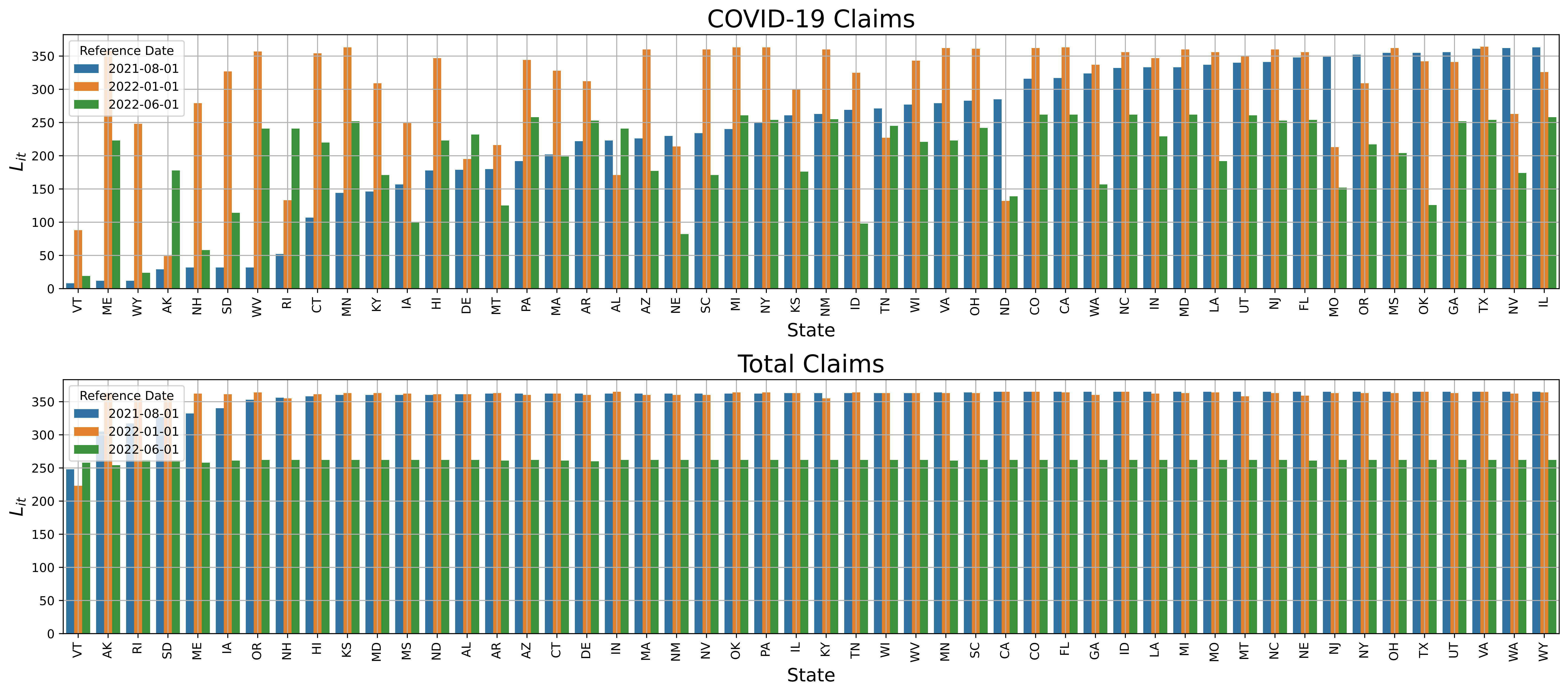

### S2_Fig

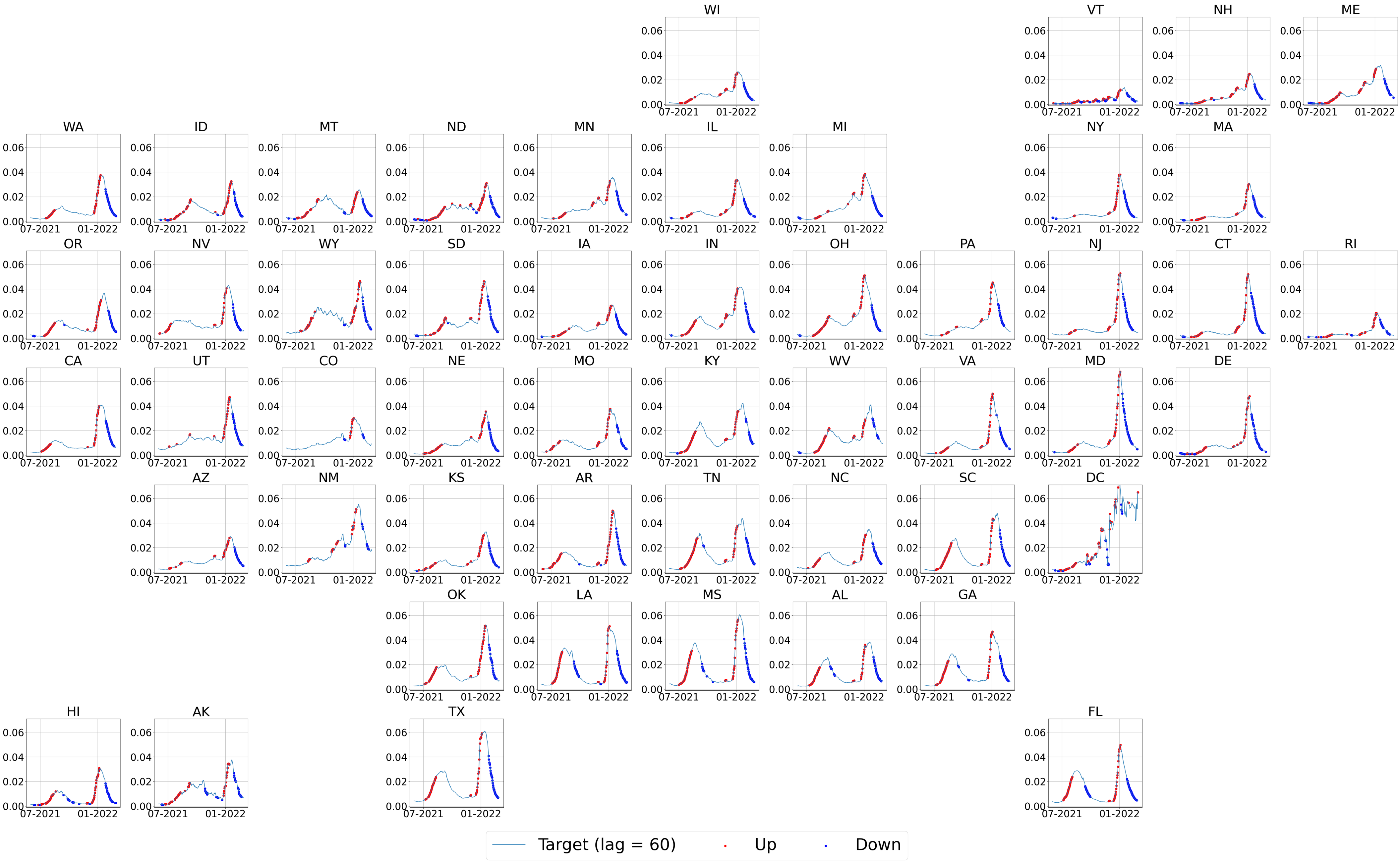

### S3_Fig

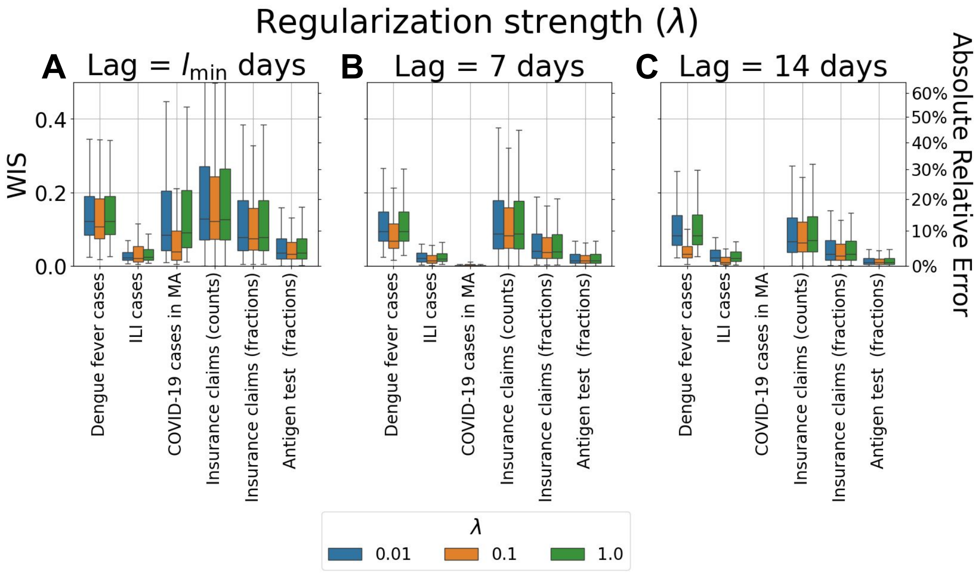

### S4_Fig

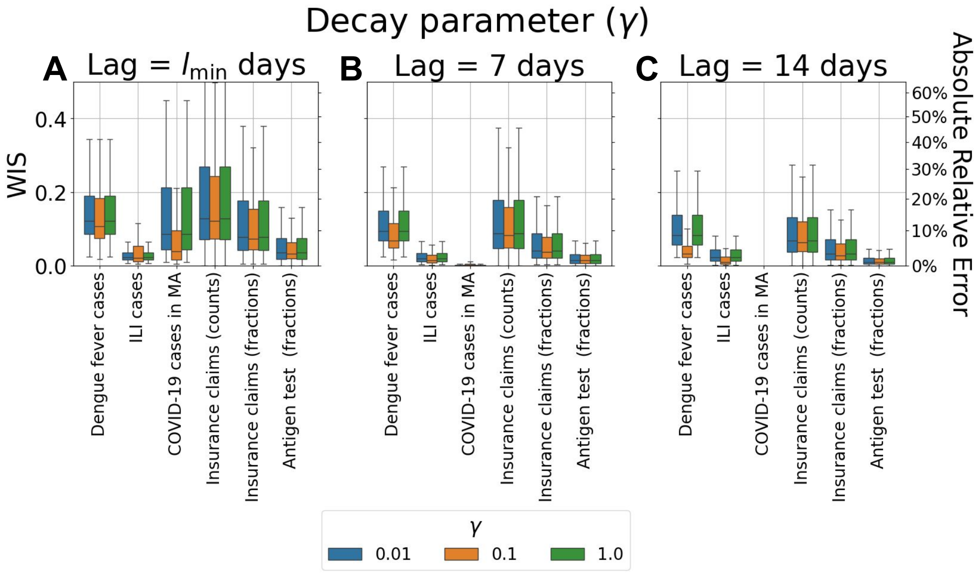

### S5_Fig

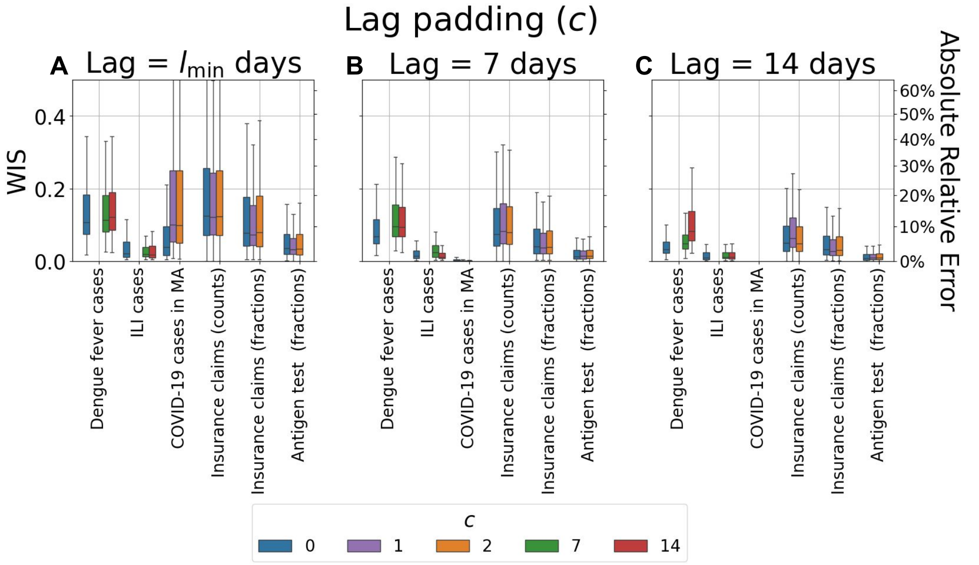
